# Glucagon/GLP-1 receptor co-agonist NNC9204-1177 reduced body weight in adults with overweight or obesity but was associated with safety issues

**DOI:** 10.1101/2022.06.02.22275920

**Authors:** Martin Friedrichsen, Lars Endahl, Frederik Flindt Kreiner, Ronald Goldwater, Martin Kankam, Søren Toubro, Sune Boris Nygård

**Affiliations:** Novo Nordisk A/S, Søborg, Denmark; PAREXEL International, Baltimore, MD, USA; Altasciences Clinical Kansas, Overland Park, KS, USA

## Abstract

Glucagon/glucagon-like peptide-1 (GLP-1) receptor co-agonists may provide greater weight loss than agonists targeting the GLP-1 receptor alone. We report results from three phase 1 trials investigating the glucagon/GLP-1 receptor co-agonist NNC9204-1177 (NN1177) for once-weekly subcutaneous use in adults with overweight or obesity.

Our focus was a 12–week multiple ascending dose (MAD), placebo-controlled, double-blind trial in which adults (N=99) received NN1177 (dose-escalated to treatment doses of 200, 600, 1,300, 1,900, 2,800, 4,200, and 6,000 μg) or placebo. Two other trials also contribute to the findings in this report: a first human dose (FHD) / single ascending dose (SAD), placebo-controlled, double-blind trial in which adults (N=49) received NN1177 (treatment doses of 10, 40, 120, 350, 700 and 1,100 μg) or placebo, and a drug–drug interaction (DDI), open-label, single-sequence trial in which adults (N=45) received a 4,200 μg dose of NN1177. Pharmacokinetic, safety and tolerability, and pharmacodynamic endpoints were assessed.

For the MAD and FHD/SAD trials, baseline characteristics were generally balanced across groups. The half-life of NN1177 was estimated at between 77.3 and 111 hours. NN1177 appeared tolerable across trials; however, a number of safety concerns were observed, including an increase in heart rate (range 5–22 beats per minute) and decrease in reticulocyte count, which were both dose dependent, and increased markers of inflammation (fibrinogen and C-reactive protein), hepatic disturbances (increased aspartate aminotransferase and alanine aminotransferase), impaired glucose tolerance (dose groups 2,800–6,000 ug) and reduced blood levels of some amino acids. Clinically relevant weight loss was achieved (up to 12.6% at week 12; 4,200 ug in the MAD trial), but this was not accompanied by cardiometabolic improvements.

In conclusion, although treatment with NN1177 was associated with dose-dependent and clinically relevant weight loss, unacceptable safety concerns precluded further clinical development.

## Introduction

Addressing the obesity pandemic is one of the largest public health challenges worldwide [1, 2]. As a chronic disease, obesity aggravates cardiometabolic risk factors such as insulin resistance, hypertension, and dyslipidaemia [3], and is associated with, for example, type 2 diabetes and cardiovascular disease [1, 4, 5]. Ultimately, obesity confers an increased risk of premature mortality [1, 6]. While a 5–10% weight loss has been shown to reduce obesity-related complications in individuals with overweight or obesity, a weight loss of at least 15% has been shown to further improve risk factors for cardiometabolic disease and improve health-related quality of life [7, 8].

Glucagon, a polypeptide structurally related to glucagon-like peptide-1 (GLP-1), and secreted by the alpha cells of the pancreas, acutely increases blood glucose through stimulation of glycogenolysis and gluconeogenesis [9]. The chronic effects of glucagon include decreased appetite combined with a possible increase in thermogenesis leading to weight loss, and an increase in lipolysis [10, 11]. Wasting, resulting from gluconeogenesis based on catabolism has also been reported in patients with glucagonomas [12, 13]. The mechanism involved in glucagon-associated appetite suppression remains under debate; however, the GLP-1 receptor (GLP-1R) could potentially be involved [14].

Glucagon-like peptide-1 is an incretin peptide hormone that stimulates insulin secretion in a glucose-dependent manner, and supresses glucagon secretion [15]. GLP-1 reduces body weight by decreasing energy intake through a reduction in appetite and induction of satiety [15]. As such, GLP-1 receptor agonists (GLP-1 RAs) have proven benefits on body weight and the complications associated with obesity [2, 16, 17]. GLP-1 RAs have been shown to reduce body weight [18, 19], and two members of the drug class, liraglutide and semaglutide, are approved for weight management. Furthermore, in type 2 diabetes, GLP-1 RAs including dulaglutide, liraglutide and semaglutide are indicated to reduce cardiovascular risk, and the cardiovascular safety of all marketed GLP-1 RAs has been confirmed [20].

Although glucagon receptor agonism increases blood glucose, simultaneous GLP-1 receptor agonism may counteract this effect, as seen when glucagon and GLP-1 are co-administered [21]. Hence, co-agonists for the glucagon and GLP-1 receptors have been shown to improve glucose tolerance and increase weight loss, compared with GLP-1 RA alone, without detrimental effects on glycaemic control [22-26]. Thus, the glucagon/GLP-1 receptor co-agonist, NNC9204-1177 (hereafter NN1177), was developed as a novel pharmacotherapeutic option for weight management to allow individuals with obesity to achieve meaningful and sustained weight loss. Of note, the preclinical evaluation of NN1177 met translational challenges, as reported separately [27].

Here, we report on three phase 1 trials investigating the safety, tolerability, pharmacokinetics (PK) and pharmacodynamics (PD) of NN1177 given as subcutaneous (s.c.) once-weekly injections to participants living with overweight or obesity. This report is based on a 12-week multiple ascending dose (MAD) trial (ClinicalTrials.gov ID NCT03308721) and clinically relevant findings from a first human dose (FHD)/single ascending dose (SAD) trial (ClinicalTrials.gov ID NCT02941042) and a drug–drug-interaction (DDI) trial (ClinicalTrials.gov NCT04059367). The focus in this report is the MAD trial, which is described in detail. The FHD/SAD trial investigated the safety, tolerability and PK of NN1177, and explored its PD properties after s.c. dosing in male participants who were overweight or with obesity, employing a sequential ascending dose design. In the MAD trial, a dose-dependent decrease in body weight from baseline was observed at the highest dose levels (600 μg to 4,200 μg) 7 days after dosing, which was not sustained during the follow-up period of 39 days post-dose. The maximum serum concentration of NN1177 (C_max_) and the area under the concentration–time curve from time zero to infinity (AUC_0-inf_) increased proportionally with increasing dose; however, because of gastrointestinal (GI) tolerability issues with the higher NN1177 dose levels, NN1177 was not tested at higher dose levels (1,600 μg and 2,000 μg). Results from the DDI trial that are relevant for the overall safety evaluation of NN1177 are reported in this article, while the main results of this trial are reported separately [28].

Briefly, the DDI trial investigated whether steady-state exposure of NN1177 after s.c. once-weekly injections influenced exposure of representative index substrates of cytochrome P450 enzymes (CYP3A4, CYP2C9, CYP2C19, CYP2D6, CYP1A2) in participants with overweight. The analysis of PK profiles showed no significant effect from the co-administration of NN1177 on the area under the concentration–time curve from time zero to infinity (AUC_0-inf_) of CYP3A4 or CYP2C9. The remaining CYP substrates generally displayed a decrease in AUC_0-inf_ and the maximum serum concentration of NN1177 (C_max_) [28]. The rationale for initiating the DDI trial was based on *in vitro* observations in which down-regulation of CYP3A4, CYP2B6, and CYP1A2 was observed at clinically relevant concentrations of NN1177.

Overall, this article presents the learnings from three clinical phase 1 trials investigating NN1177. Although clinical development of the glucagon/GLP-1 receptor co-agonist was discontinued, the results and interpretations represent relevant challenges and mechanisms potentially associated with the clinical development and use of this drug class.

## Research Design and Methods

Three phase 1 trials are described herein: (1) the MAD trial (primary focus of this manuscript); (2) the FHD/SAD trial (to fully characterise the compound, complement MAD trial data on safety endpoints of specific interest and body weight results); and (3) the DDI trial (to complement safety endpoints of specific interest the main results of this trial will be reported elsewhere in a separate publication currently under review) [28].

### Trial designs

Trial designs for the MAD and FHD/SAD trials and a full list of inclusion/exclusion criteria are shown in Supplementary Appendix S1 (Protocol NNC9277–4328, section 6.2, page 39 for the MAD trial and Protocol NN9277–4258, section 6.2, page 33 for the FHD/SAD trial). The studies obtained approval from local institutional review boards with written consent obtained from all participants.

#### Multiple ascending dose trial

The MAD trial (NCT03308721) was a single-centre, placebo-controlled, double-blind, randomised, sequential ascending multiple-dose clinical trial. Seven cohorts of adult participants (age 18–55 years) with overweight or obesity (body mass index [BMI] 27.0–39.9 kg/m^2^) were randomised (9:3) to receive ascending multiple doses of once-weekly s.c. NN1177 or placebo. Exclusion criteria included: age ≥40 years with an estimated ≥5% risk of 10-year atherosclerotic disease. Seven dose levels (200, 600, 1,300, 1,900, 2,800, 4,200, and 6,000 μg) of NN1177 were investigated in ascending order. In each cohort, a total of 12 participants were treated for up to 12 weeks. To mitigate GI adverse effects associated with GLP-1 RA treatment, dose escalation to the final dose level was applied in cohorts two to seven (Supplementary Table S1). In-house safety monitoring periods were planned approximately every second week and coincided with dose escalation visits in cohorts two to seven. Between cohorts, safety parameters were reviewed to decide whether to ascend to the next dose level. Protocol-defined subject-level stopping rules were plasma glucose ≥11.1 mM (200 mg/dL) during a 2-hour oral glucose tolerance test (OGTT) or cardiovascular abnormalities measured after at least 10 minutes rest in the supine position. The specific cardiovascular parameters were QT interval prolongation (increase from baseline of >60 ms for the heart-rate corrected interval [QT corrected for heart rate by Fridericia’s cube root formula; QTcF]), or absolute heart rate of >115 beats per minute (bpm) or sustained absolute heart rate of >100 bpm over 24 hours. Participants were initially treated on day 1 (baseline), continued treatment until day 85 (end of treatment) and were followed up for safety until day 110.

#### First human dose/single ascending dose trial

The FHD/SAD trial (NCT02941042) was a single-centre, placebo-controlled, double-blind, randomised, single-dose clinical trial. Exclusion criteria included: estimated ≥5% risk of 10-year atherosclerotic disease; history or clinically relevant conditions (including diabetes); prior obesity surgery; abnormal laboratory or electrocardiography results. Six cohorts of male adult participants (age 18–55 years) with overweight or obesity (BMI 25.0–34.9 kg/m^2^) were randomised (3:1) to receive ascending single doses of s.c. NN1177 or placebo. Eight cohorts (N=8, including two participants receiving placebo) receiving s.c. NN1177 (10, 40, 120, 350, 700, 1,100 μg) were planned. Participants were followed for 39 days post-dose.

#### Drug-drug interaction trial

The DDI trial is described in detail elsewhere (manuscript under development) [28, 29]; in brief, this was a single centre, open-label, single-sequence trial. Adult participants (age 18–60 years) with overweight (BMI 23.0–29.9 kg/m^2^ and HbA_1c_ <6.5%) received once-weekly s.c. NN1177 (dose escalated every 2 weeks until a final dose of 4,200 μg for 3 weeks). NN1177 was initiated on day 1 (baseline) and the last dose was administered on day 78 (end of treatment). A Cooperstown 5+1 index cocktail, containing representative index substrates for multiple CYP enzymes, was administered before NN1177 treatment on day 1, and on day 78 prior to the final 4,200 μg dose of NN1177.

### Endpoints

#### Pharmacokinetic endpoints

PK characteristics of NN1177 were assessed as supportive secondary endpoints in both the MAD and the FHD/SAD trials (assessed at steady state in the MAD trial and after single dose in the FHD/SAD trial). The PK endpoints included the area under the NN1177 serum concentration–time curve from time 0 to 168 h (AUC_0-168h_), C_max_, the time to maximum serum concentration of NN1177 (t_max_) and the terminal half-life (t_½_). Exploratory PK endpoints assessed in both trials (at steady state in the MAD trial and after single dose in the FHD/SAD trial) comprised the apparent total serum clearance of NN1177 (CL/F), the apparent volume of distribution (at steady state and during elimination in the MAD trial (V_z_/F) and the mean residence time (MRT). PK parameters assessed in the DDI trial are not reported herein.

#### Safety and tolerability endpoints (including primary endpoint)

For both the MAD and FHD/SAD trials, the primary endpoint was the number of all treatment-emergent adverse events (TEAEs) recorded from time of dosing of NN1177 until completion of the post-treatment follow-up visit. TEAEs were also recorded in the DDI trial, and safety endpoints of specific interest are reported herein. As well as all TEAEs, injection-site reactions were assessed at prespecified time points (MAD trial). Details of exploratory endpoints in the MAD and FHD/SAD trials are found in the protocols supplied in Supplementary Appendix 1 (Protocol NN9277-4328, section 4.2.3, page 30 and Protocol NN9277-4258, section 4.2.2, page 24).

The potential exposure–response relationship of NN1177 and changes in the QTc interval obtained from Holter monitoring was also evaluated in the MAD trial. Prior to database lock, the prespecified dependent variable QTcF was changed to individual-specific corrected QT interval (QTcI) to adequately correct for the increased heart rate.

#### Pharmacodynamic endpoints

Change in body weight was assessed from baseline to end of treatment as an exploratory PD endpoint in the MAD trial, and from baseline to end of follow-up as a supportive secondary PD endpoint in the FHD/SAD trial. Glucose metabolism parameters were also assessed as exploratory PD endpoints, including glucose metabolism parameters (fasting glucose, fasting insulin, fasting glucagon and HbA_1c_) and hormones (fasting leptin and fasting soluble leptin receptor) which were assessed from baseline to end of treatment in the MAD trial. An OGTT was conducted on day 1, day 29 and day 85 in the MAD trial. For each OGTT, the following endpoints were assessed: area under the curve from time 0 to 2 h (AUC_0-2h_) for insulin and glucose, incremental AUC from time 0 to 2 h (iAUC_0-2h /2h_) for insulin and glucose, insulinogenic index (the increase in insulin [0–30 minutes]/increase in glucose [0−30 minutes]), insulin secretion ratio (the ratio of the AUCs calculated from time 0 to 2 h during OGTT for insulin and glucose). In addition, blood samples for assessment of glucose (measured in plasma) and insulin were taken 0, 10, 20, 30, 60, 90 and 120 minutes after glucose intake.

### Statistical analysis

For both the MAD and the FHD/SAD trials, analyses of safety endpoints used the safety analysis set consisting of all participants who were exposed to at least one dose of NN1177. Analyses of PK and PD endpoints were based on the full analysis set consisting of all participants who were randomised and received at least one dose of NN1177. PK endpoints were compared between treatments using an analysis of variance (ANOVA) model based on log-transformed endpoints with dose as a factor. Dose proportionality analysis of PK endpoints was performed using a linear regression model based on log-transformed PK endpoints, log(dose) as covariate. QTcI exposure–response analysis was assessed using a mixed-effect model for repeated measurements on change from baseline in QTcI, with fixed effects for treatment (active/placebo), time point, baseline QTcI and NN1177 concentration and random effects for subject and NN1177 concentration [30]. No methods for imputing missing data were applied.

## Results

### Dose levels achieved

Dosing was conducted up to 6,000 μg in the MAD trial and up to 1,100 μg in the FHD/SAD trial. As per the trial protocol, a trial safety group in both trials blind-reviewed preliminary safety, PK and PD data before dose escalation to higher, never-tested plasma exposures. In the MAD trial, dose escalation to the top dose level of 6,000 μg in cohort seven was modified on the basis of data from cohort one to six; the regimen was modified to incorporate a lower initial dose level (Supplementary Table S1). In the FHD/SAD trial, GI tolerability concerns (based on review by the trial safety group of the frequency and severity of events [preferred terms] within the ‘GI disorder’ system organ class) were raised following administration of 1,100 μg. Consequently, it was decided not to initiate the planned 1,600 μg and 2,000 μg cohorts in the FHD/SAD trial.

### Subject disposition, baseline characteristics and dose levels

Participant flows are shown in Figure 1 for the MAD trial (Figure 1A) and FHD/SAD trial (Figure 1B).

**Figure 1.**
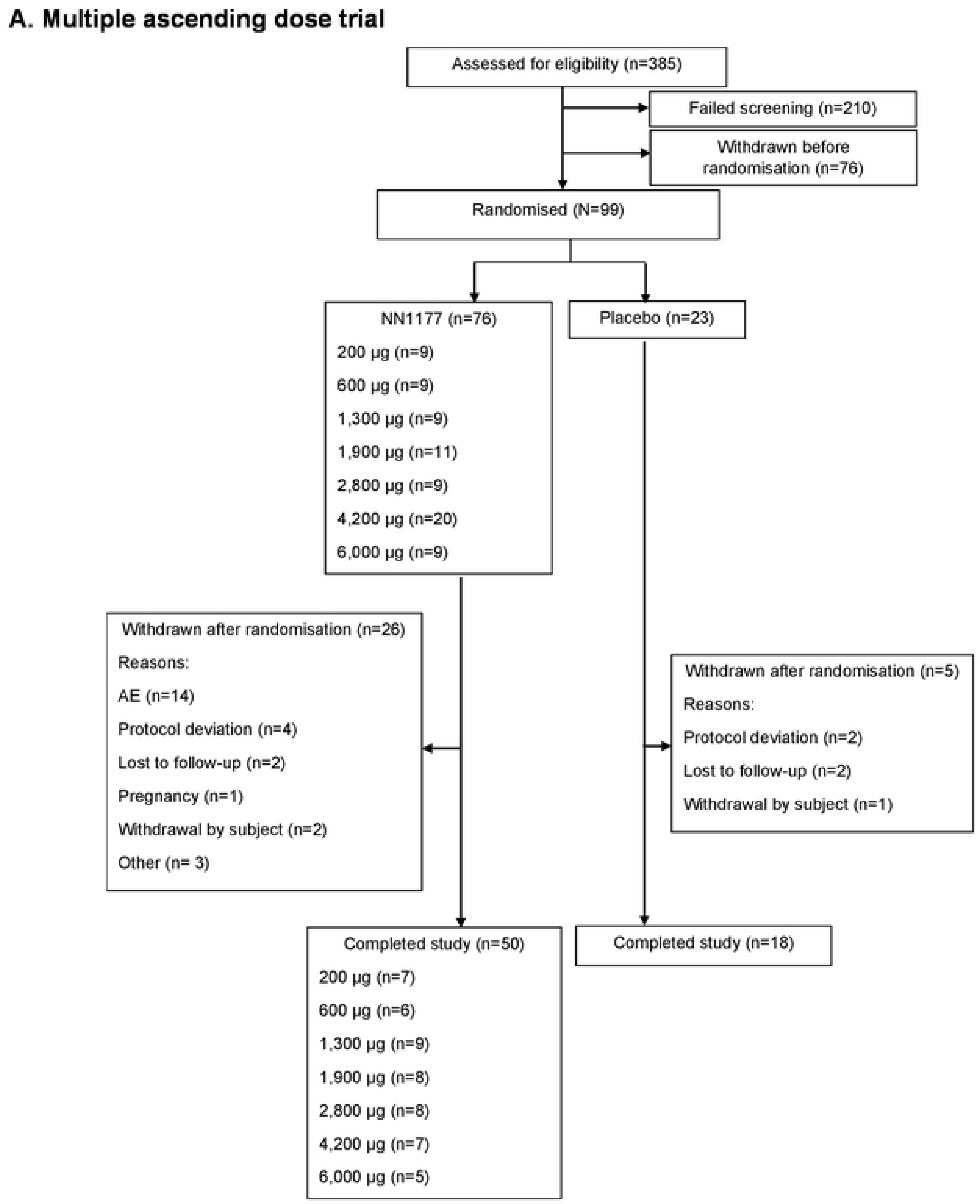
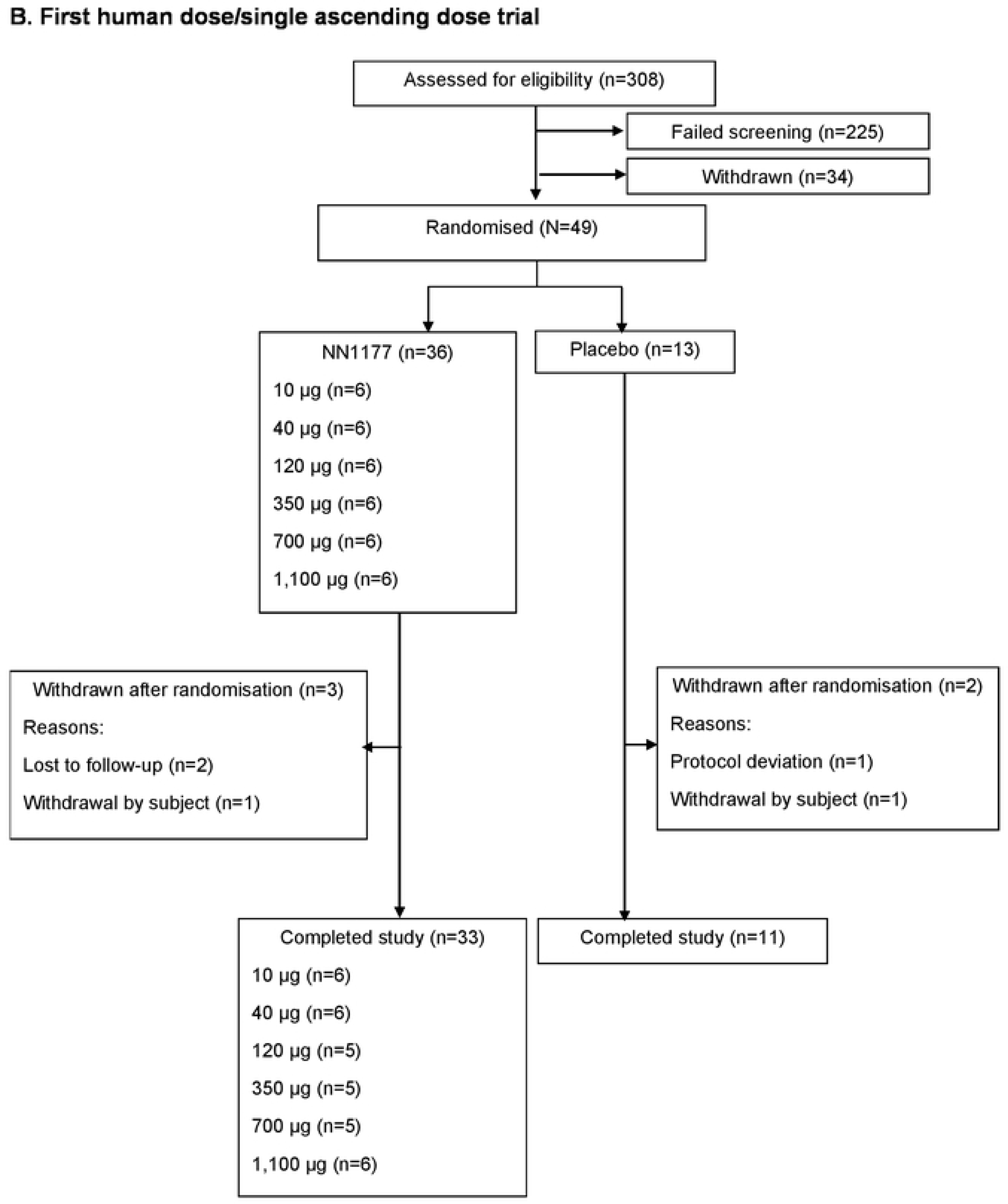
Participant flow. **A. Multiple ascending dose trial** The number of participants exposed to NN1177 1,900 μg (n=11) and 4,200 μg (n=20) was higher compared with those of the other NN1177 groups. In cohort 4 (1,900 μg) and cohort 6 (4,200 μg), a blinded assessment of the timing and reasons for participants being withdrawn after randomisation led to replacements of participants in order to attain at least 8 participants for safety evaluation. AE, adverse event; n, number of participants. **B. First human dose/single ascending dose trial** AE, adverse event; n, number of participants

In the MAD trial, 99 participants were randomised and exposed to active treatment (200 μg, 600 μg, 1,300 μg [n=9 each], 1,900 μg [n=11], 2,800 μg [n=9], 4,200 μg [n=20], and 6,000 μg [n=9]) or placebo (n=23). More participants were randomised and exposed to the 1,900 μg (n=11) and 4,200 μg (n=20) dose levels compared with other dose levels (n=9). This was due to a higher number of withdrawals substituted to achieve at least eight participants for safety evaluation. A total of 68 participants (68.7%) completed the trial. More participants withdrew after randomisation in the NN1177 4,200 μg group (13 participants [65.0%]) than in the other NN1177 groups (Supplementary Appendix S2). In the FHD/SAD trial, 49 participants were randomised and received NN1177. Five participants were withdrawn after randomisation (Supplementary Appendix S2), with 44 participants (89.8%) completing the trial. In the DDI trial, 45 participants were enrolled, of whom 37 completed the trial.

Table 1 shows baseline characteristics for the MAD trial and FHD/SAD trial, with the baseline characteristics for the DDI trial shown in Supplementary Table S2.

**Table 1.**
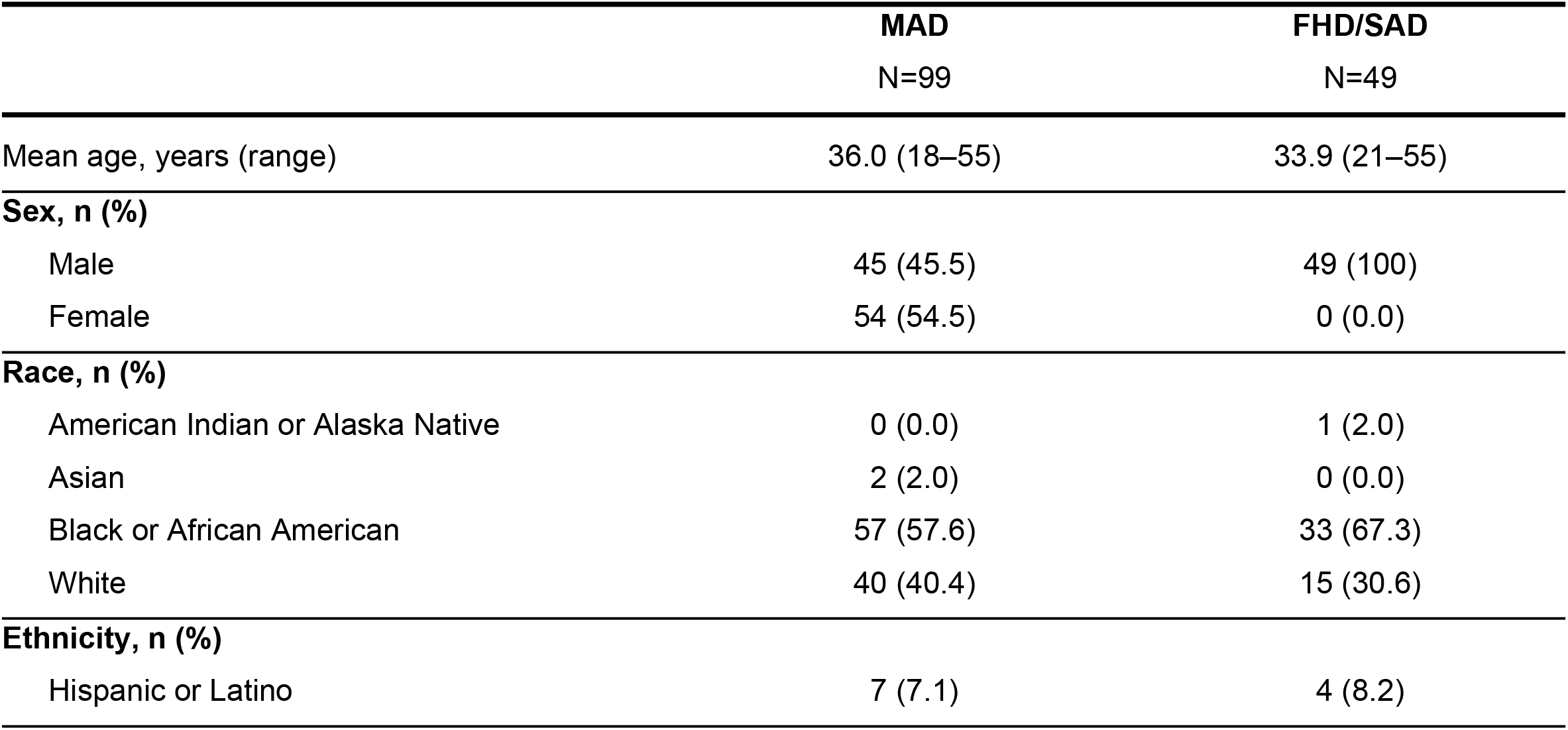
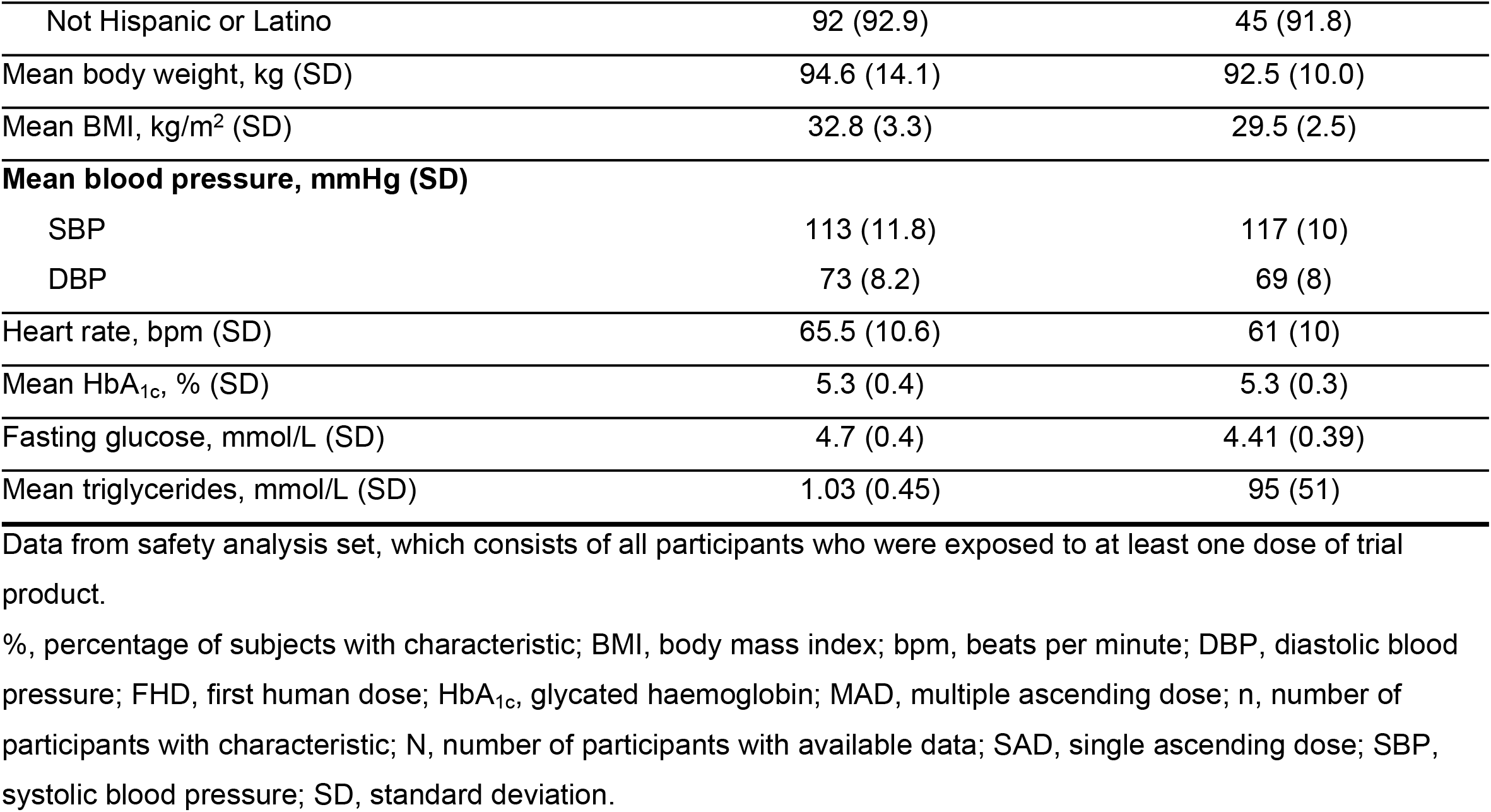
Baseline characteristics in the multiple ascending dose and first human dose/single ascending dose trials.

|  | **MAD**<br>N=99 | **FHD/SAD**<br>N=49 |
| --- | --- | --- |
| Mean age, years (range) | 36.0 (18–55) | 33.9 (21–55) |
| **Sex, n (%)** |  |  |
| Male | 45 (45.5) | 49 (100) |
| Female | 54 (54.5) | 0 (0.0) |
| **Race, n (%)** |  |  |
| American Indian or Alaska Native | 0 (0.0) | 1 (2.0) |
| Asian | 2 (2.0) | 0 (0.0) |
| Black or African American | 57 (57.6) | 33 (67.3) |
| White | 40 (40.4) | 15 (30.6) |
| **Ethnicity, n (%)** |  |  |
| Hispanic or Latino | 7 (7.1) | 4 (8.2) |
| Not Hispanic or Latino | 92 (92.9) | 45 (91.8) |
| Mean body weight, kg (SD) | 94.6 (14.1) | 92.5 (10.0) |
| Mean BMI, kg/m<sup>2</sup> (SD) | 32.8 (3.3) | 29.5 (2.5) |
| **Mean blood pressure, mmHg (SD)** |  |  |
| SBP | 113 (11.8) | 117 (10) |
| DBP | 73 (8.2) | 69 (8) |
| Heart rate, bpm (SD) | 65.5 (10.6) | 61 (10) |
| Mean HbA<sub>1c</sub>, % (SD) | 5.3 (0.4) | 5.3 (0.3) |
| Fasting glucose, mmol/L (SD) | 4.7 (0.4) | 4.41 (0.39) |
| Mean triglycerides, mmol/L (SD) | 1.03 (0.45) | 95 (51) |

Data from safety analysis set, which consists of all participants who were exposed to at least one dose of trial product.
%, percentage of subjects with characteristic; BMI, body mass index; bpm, beats per minute; DBP, diastolic blood pressure; FHD, first human dose; HbA<sub>1c</sub>, glycated haemoglobin; MAD, multiple ascending dose; n, number of participants with characteristic; N, number of participants with available data; SAD, single ascending dose; SBP, systolic blood pressure; SD, standard deviation.

For the MAD and FHD/SAD trials, demographics and baseline characteristics were generally similar across treatment groups (Supplementary Table S3). There were no clinically relevant imbalances across treatment groups with respect to medical history at screening.

### Pharmacokinetic characterisation

NN1177 serum concentration profiles for all doses in the MAD and FHD/SAD trials are presented in Supplementary Figure S1. AUC_0-∞_ and C_max_ at steady state (AUC_0-∞,ss_ and C_max,ss_, respectively) were consistent with dose proportionality in the FHD trial. In the MAD trial, although mean NN1177 serum concentrations appeared to increase with increasing dose, a minor deviance from dose proportionality was observed in steady-state PK (Table 2). The median t_max,_ at steady state (t_max,SS_) ranged from 30 to 48 hours in the MAD trial and from 33 to 45 hours in the FHD/SAD trial (Table 2 and Supplementary Table S4). The geometric mean t_1/2,_ at steady state (t_1/2,SS_) ranged from 77.3 to 111 hours for the MAD trial (Table 2) and the geometric mean of t_1/2,SS_ ranged from 76.2 to 87.0 hours for the FHD/SAD trial with no apparent dose relationship (Supplementary Table S4).

**Table 2.** Pharmacokinetic endpoints in the multiple ascending dose trial.

|  | NN1177<br>200 µg | NN1177<br>600 µg | NN1177<br>1,300 µg | NN1177<br>1,900 µg | NN1177<br>2,800 µg | NN1177<br>4,200 µg | NN1177<br>6,000 µg |
| --- | --- | --- | --- | --- | --- | --- | --- |
| **AUC<sub>0-168h,SS</sub> (nmol\*h/L)\*** |  |  |  |  |  |  |  |
| Geometric mean (CV) | 559 (28.8) | 1,729 (31.7) | 4,334 (34.8) | 7,586 (24.8) | 11,299 (37.2) | 17,197 (35.1) | 27,959 (31.3) |
| Min.; Max. | 379; 871 | 1,273; 3,101 | 2,491; 8,114 | 5,450; 10,723 | 7,637; 19,334 | 12,097; 34,192 | 19,561; 43,425 |
| **C<sub>max,SS</sub> (nmol/L)\*** |  |  |  |  |  |  |  |
| Geometric mean (CV) | 4.63 (27.4) | 14.1 (40.1) | 33.3 (36.9) | 60.5 (26.0) | 87.6 (38.3) | 132 (36.5) | 239 (30.0) |
| Min.; Max. | 3.24; 6.87 | 10.5; 29.9 | 17.9; 60.7 | 40.1; 83.2 | 59.1; 150 | 89.1; 255 | 185; 389 |
| **t<sub>1/2,SS</sub> (h)** |  |  |  |  |  |  |  |
| Geometric mean (CV) | 77.3 (10.3) | 104 (40.0) | 94.6 (20.8) | 93.3 (13.5) | 101 (26.7) | 111 (14.2) | 92.7 (25.0) |
| Min.; Max. | 68.0; 92.2 | 66.4; 184 | 71.6; 131 | 79.3; 123 | 787; 169 | 97.5; 148 | 71.2; 133 |
| **CL/F<sub>SS</sub> (L/h)** |  |  |  |  |  |  |  |
| Geometric mean (CV) | 0.0783 (28.8) | 0.0759 (31.7) | 0.0656 (34.8) | 0.0548 (24.8) | 0.0542 (37.2) | 0.0534 (35.1) | 0.0470 (31.3) |
| Min.; Max. | 0.0502; 0.116 | 0.0423; 0.103 | 0.0351; 0.114 | 0.0388; 0.0763 | 0.0317; 0.0802 | 0.0269; 0.0760 | 0.0302; 0.0671 |
| **t<sub>max,SS</sub> (h)** |  |  |  |  |  |  |  |
| Median | 30 | 33 | 48 | 48 | 42 | 39 | 48 |
| Min.; Max. | 24; 48 | 12; 48 | 12; 72 | 30; 72 | 12; 48 | 24; 48 | 48; 48 |
| **MRT (h)** |  |  |  |  |  |  |  |
| Geometric mean (CV) | 138 (9.0) | 169 (16.1) | 172 (16.1) | 161 (10.0) | 189 (25.4) | 204 (7.1) | 167 (20.3) |
| Min.; Max. | 128; 159 | 132; 210 | 140; 221 | 132; 176 | 143; 316 | 189; 236 | 129; 225 |

Data from full analysis set consisting of all participants who were randomised and received at least one dose of trial product. Blood samples were drawn for bioanalysis of serum concentrations of NN1177 from day 78 until the follow-up visit (day 110). \*2<sup>β</sup> =2 corresponds to dose proportionality, therefore, a minor deviance from dose proportionality was observed as follows; AUC<sub>0-168h,SS</sub> [estimate for 2<sup>β</sup>: 2.23 nmol\*h/L [95% CI 2.10–2.36], C<sub>max,SS</sub> [estimate for 2<sup>β</sup>: 2.21 nmol/L [95% CI 2.08;2.35]; AUC<sub>0-168h,SS</sub>, area under the NN1177 serum concentration–time curve from time 0 to 168 h at steady state; CI, confidence interval; CL/F<sub>SS</sub>, apparent total serum clearance of NN1177 at steady state; C<sub>max,SS</sub>, maximum serum concentration of NN1177 at steady state; CV, coefficient of variation; h, hours; MAD, multiple ascending dose; Max., maximum; Min., minimum; MRT, mean residence time; PK, pharmacokinetic; t<sub>1/2,SS</sub>, terminal half-life at steady state; t<sub>max,SS</sub>, time to maximum serum concentration of NN1177 at steady state.

### Safety and tolerability

#### General safety and tolerability

In the MAD trial, 94.7% (n=72; 594 events) of participants reported TEAEs with NN1177 compared with 69.6% (n=16; 62 events) with placebo (Table 3). Generally, fewer participants in dose groups 200 μg and 600 μg reported TEAEs compared with those receiving the higher doses, but there was no clear dose relationship (Table 3). Across groups with a tendency towards a dose relationship for NN1177 (≥1,300 μg dose groups), most TEAEs were of mild or moderate severity and most participants had TEAEs that were judged by the investigator as probably or possibly related to the NN1177. Amongst participants receiving NN1177, 13 participants (13 events) withdrew from the trial due to a TEAE, compared with no participants in the placebo group (Table 3). Five participants receiving NN1177 withdrew from the trial because they met a stopping rule criterion: 3 participants met the heart rate criterion (n=1 in the 1,900 μg dose group [>100 bpm] and n=2 in the 4,200 μg dose group [one at >100 bpm and one at >115 bpm]); 2 participants met the OGTT criterion (n=1 in 200 μg dose group and n=1 in 6,000 μg dose group).

**Table 3.** Treatment-emergent adverse events in the multiple ascending dose trial.

|  | NN1177<br>200 µg |  |  | NN1177<br>600 µg |  |  | NN1177<br>1,300 µg |  |  | NN1177<br>1,900 µg |  |  | NN1177<br>2,800 µg |  |  | NN1177<br>4,200 µg |  |  | NN1177<br>6,000 µg |  |  | Placebo |  |  | Total |  |  |
| --- | --- | --- | --- | --- | --- | --- | --- | --- | --- | --- | --- | --- | --- | --- | --- | --- | --- | --- | --- | --- | --- | --- | --- | --- | --- | --- | --- |
|  | n | (%) | E | n | (%) | E | n | (%) | E | n | (%) | E | n | (%) | E | n | (%) | E | n | (%) | E | n | (%) | E | n | (%) | E |
| **Number of participants** | 9 |  | 0 | 9 |  | 0 | 9 |  | 0 | 11 |  | 0 | 9 |  | 0 | 20 |  | 0 | 9 |  | 0 | 23 |  | 0 | 99 |  | 0 |
| **Adverse events** | 8 | (88.9) | 29 | 6 | (66.7) | 37 | 9 | (100) | 31 | 11 | (100) | 90 | 9 | (100) | 95 | 20 | (100) | 219 | 9 | (100) | 93 | 16 | (69.6) | 62 | 88 | (88.9) | 656 |
| Serious adverse events | 0 |  | 0 | 0 |  | 0 | 0 |  | 0 | 0 |  | 0 | 0 |  | 0 | 0 |  | 0 | 1 | (11.1) | 6 | 0 |  | 0 | 1 | (1.0) | 6 |
| Events leading to withdrawal from trial* | 1 | (11.1) | 1 | 1 | (11.1) | 1 | 0 |  | 0 | 1 | (9.1) | 1 | 1 | (11.1) | 1 | 6 | (30.0) | 6 | 3 | (33.3) | 3 | 0 |  | 0 | 13 | (13.1) | 13 |
| **Treatment-related†** |  |  |  |  |  |  |  |  |  |  |  |  |  |  |  |  |  |  |  |  |  |  |  |  |  |  |  |
| Probably | 6 | (66.7) | 16 | 3 | (33.3) | 15 | 6 | (66.7) | 21 | 11 | (100) | 60 | 9 | (100) | 65 | 19 | (95.0) | 156 | 9 | (100) | 68 | 12 | (52.5) | 31 | 75 | (75.8) | 432 |
| Possibly | 2 | (22.2) | 3 | 4 | (44.4) | 8 | 0 |  |  | 7 | (63.6) | 19 | 9 | (100) | 22 | 16 | (80.0) | 43 | 8 | (88.9) | 15 | 9 | (39.1) | 13 | 55 | (55.6) | 123 |
| Unlikely | 6 | (66.7) | 10 | 6 | (66.7) | 14 | 8 | (88.9) | 10 | 7 | (63.6) | 11 | 4 | (44.4) | 8 | 13 | (65.0) | 20 | 5 | (55.6) | 10 | 13 | (56.5) | 18 | 62 | (62.6) | 101 |
| **Severity** |  |  |  |  |  |  |  |  |  |  |  |  |  |  |  |  |  |  |  |  |  |  |  |  |  |  |  |
| Severe | 0 |  | 0 | 0 |  | 0 | 0 |  | 0 | 0 |  | 0 | 0 |  | 0 | 0 |  | 0 | 1 | (11.1) | 5 | 0 |  | 0 | 1 | (1.0) | 5 |
| Moderate | 1 | (11.1) | 2 | 4 | (44.4) | 5 | 1 | (11.1) | 1 | 3 | (27.3) | 10 | 5 | (55.6) | 9 | 8 | (40.0) | 18 | 4 | (44.4) | 16 | 3 | (13.0) | 3 | 29 | (29.3) | 64 |
| Mild | 8 | (88.9) | 27 | 6 | (66.7) | 32 | 9 | (100) | 30 | 11 | (100) | 80 | 9 | (100) | 86 | 20 | (100) | 201 | 9 | (100) | 72 | 15 | (65.2) | 59 | 87 | (87.9) | 587 |

Data from safety analysis set consisting of all participants who were exposed to at least 1 dose of trial product. TEAEs recorded from time of dosing of NN1177 (day 1) until completion of the post-treatment follow-up visit (day 110). AEs were coded using the most recent version of MedDRA (version 22.1). Participants who received placebo were pooled across cohorts.
\*The TEAEs leading to withdrawal were (doses in bold refer to the assigned dose group whereas doses not in bold refer to the dose level achieved [in escalation] when the event occurred): 13 TEAEs leading to withdrawal; **200 µg**: blood glucose increased (n=1) at dose 200 µg, **600 µg**: upper respiratory tract infection (n=1) at dose 600 µg, **1,900 µg**: vomiting (n=1) at dose 1,300 µg, **2,800 µg**: arrhythmia (n=1) at dose 1,300 µg, **4,200 µg**: abdominal pain (n=2) at doses 1,300 µg and 1,900 µg, asthenia (n=1) at dose 2,800 µg, sinus tachycardia (n=2) at doses 1,300 µg and 4,200 µg, nausea (n=1) at dose 4,200 µg, **6,000 µg**: vomiting (n=1) at dose 3,200 µg, fatigue n=(1) at dose 3,200 µg, hyperglycaemia (n=1) at dose 1,600 µg; † As assessed by investigator.
%, percentage of participants with an AE; AE, adverse events; E, number of events; MedDRA, Medical Dictionary for Regulatory Activities; n, number of participants; SAE, serious adverse event; TEAE, treatment-emergent adverse event.

Events related to GI tolerability (‘GI disorders’ system organ class), especially nausea and vomiting, were the most frequently reported TEAEs and were experienced by 58.6% of participants (202 events in 58 participants) with NN1177 and 30.4% of participants (11 events in seven participants) with placebo (Supplementary Table S5). The proportion of GI TEAEs possibly or probably related to NN1177 treatment was highest for participants treated with dose levels ≥ 1,900 μg. Injection-site reactions (pain, itching and redness) were mild, and reported by 28.3% (89 events in 28 participants) of participants with NN1177 and 26.1% (11 events in 6 participants) of participants with placebo. The AE profiles observed in the FHD/SAD and DDI trials (Supplementary Tables S6 and S7) generally reflected the profile observed in the MAD trial.

#### Heart rate, QT interval and blood pressure

Increases in heart rate were seen in all three trials. From baseline to end of treatment, heart rate increased by 1–15 bpm with NN1177 in the MAD trial (at day 85, 168 hours after last dosing; Figure 2A) and by up to 22 bpm in the FHD/SAD trial (24 hours post-dosing). In the MAD trial, heart rate increases appeared to be dose-dependent and more pronounced at the highest doses (Figure 2A). At follow-up in the MAD and FHD/SAD trials, the heart rate increases had, in general, reversed or diminished. In the DDI trial, heart rate was measured continuously over 48 hours, with observed increases greatest at night (mean of 15 bpm) and higher than average increases over 24 hours (12 bpm). The difference in average heart rate between night and day seen at baseline (day 0) diminished following treatment with NN1177 (day 65; Figure 2B). In the MAD trial, treatment with NN1177 appeared to prolong the QTcI by up to 18 msec, in a concentration-dependent manner (Figure 2C). Figures for QTcI as a function of time and model diagnostic plots are available in Supplementary Figures S3 and S4. No clinically relevant changes in systolic or diastolic blood pressure were observed in the MAD or FHD/SAD trials.

**Figure 2.**
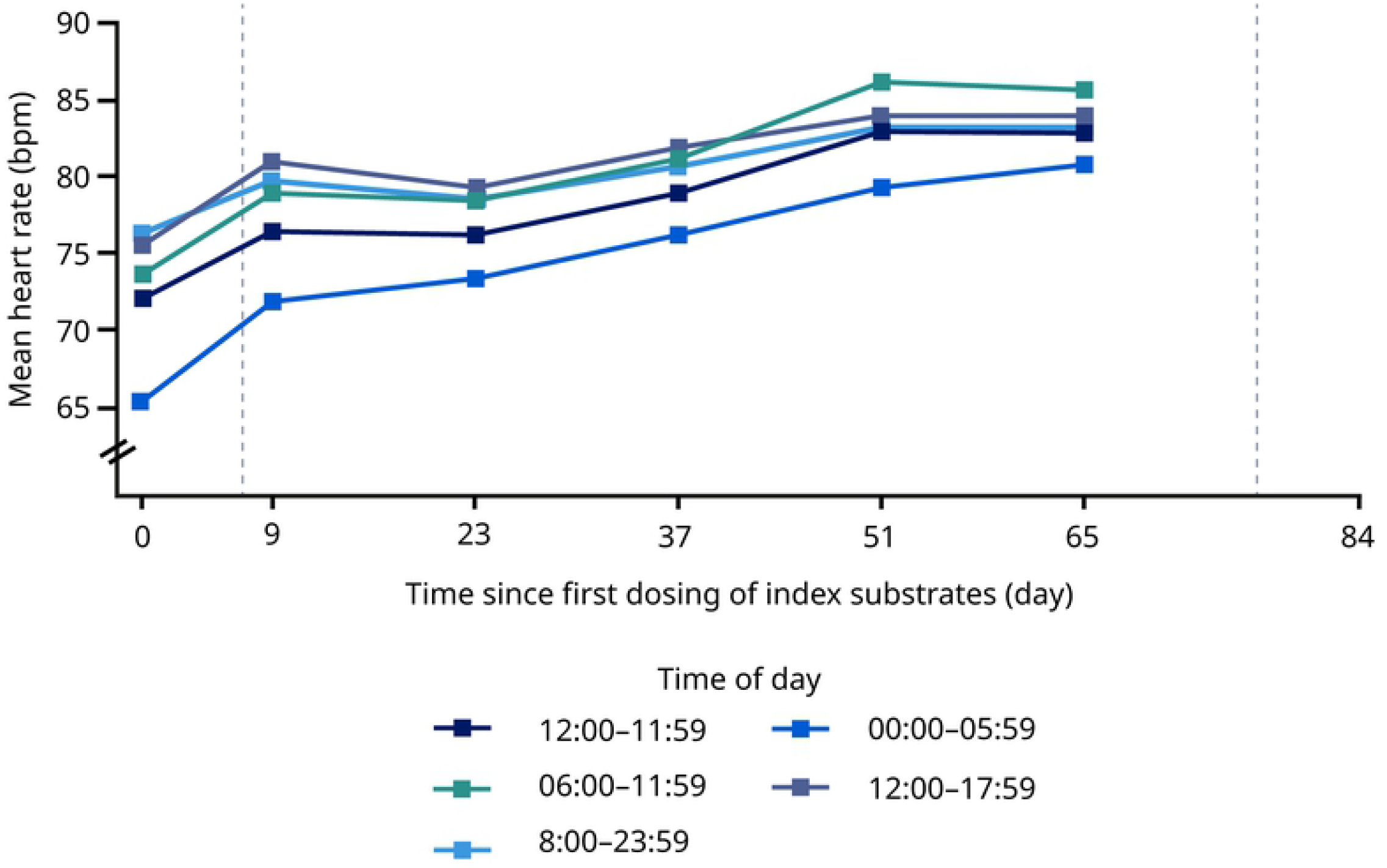
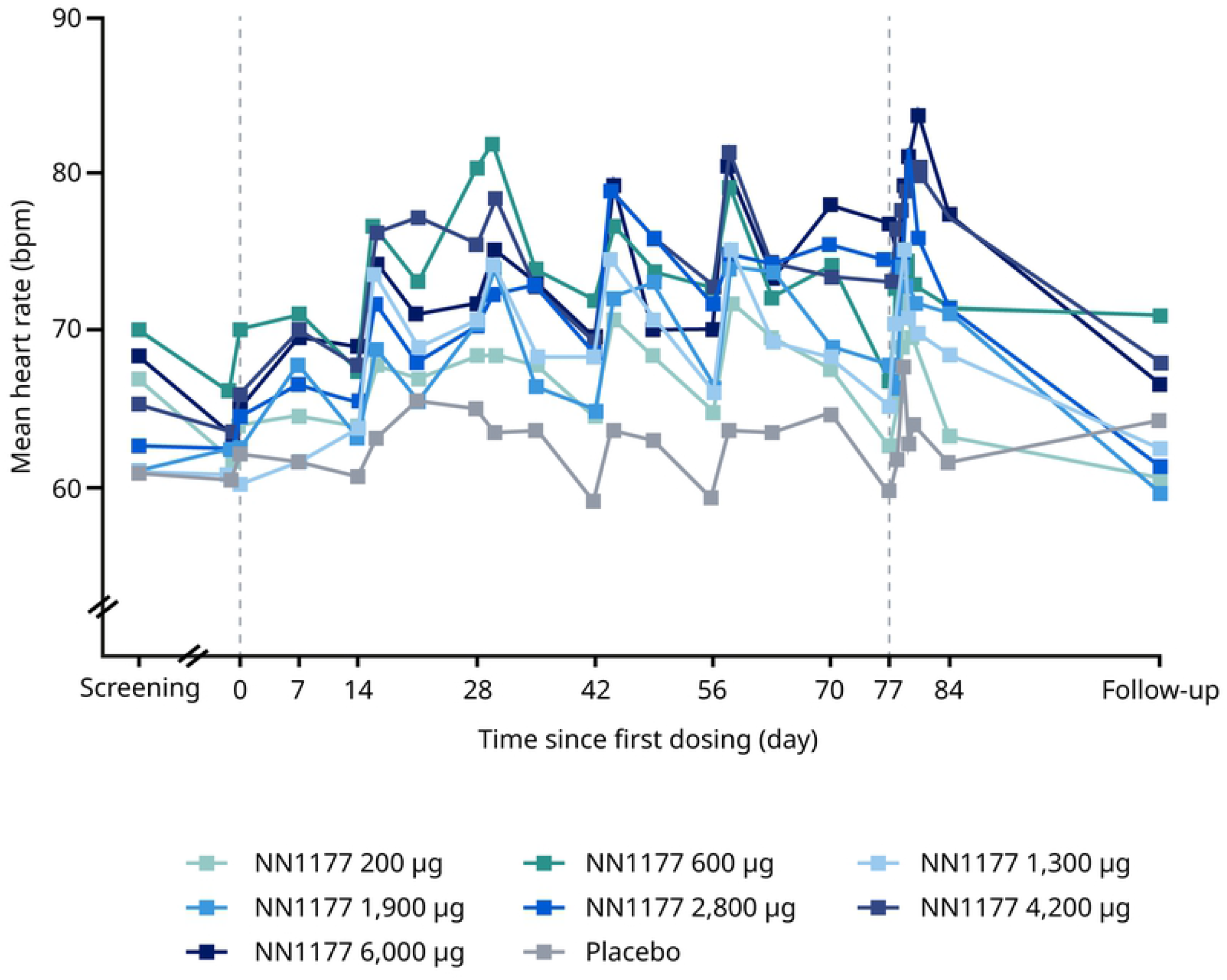
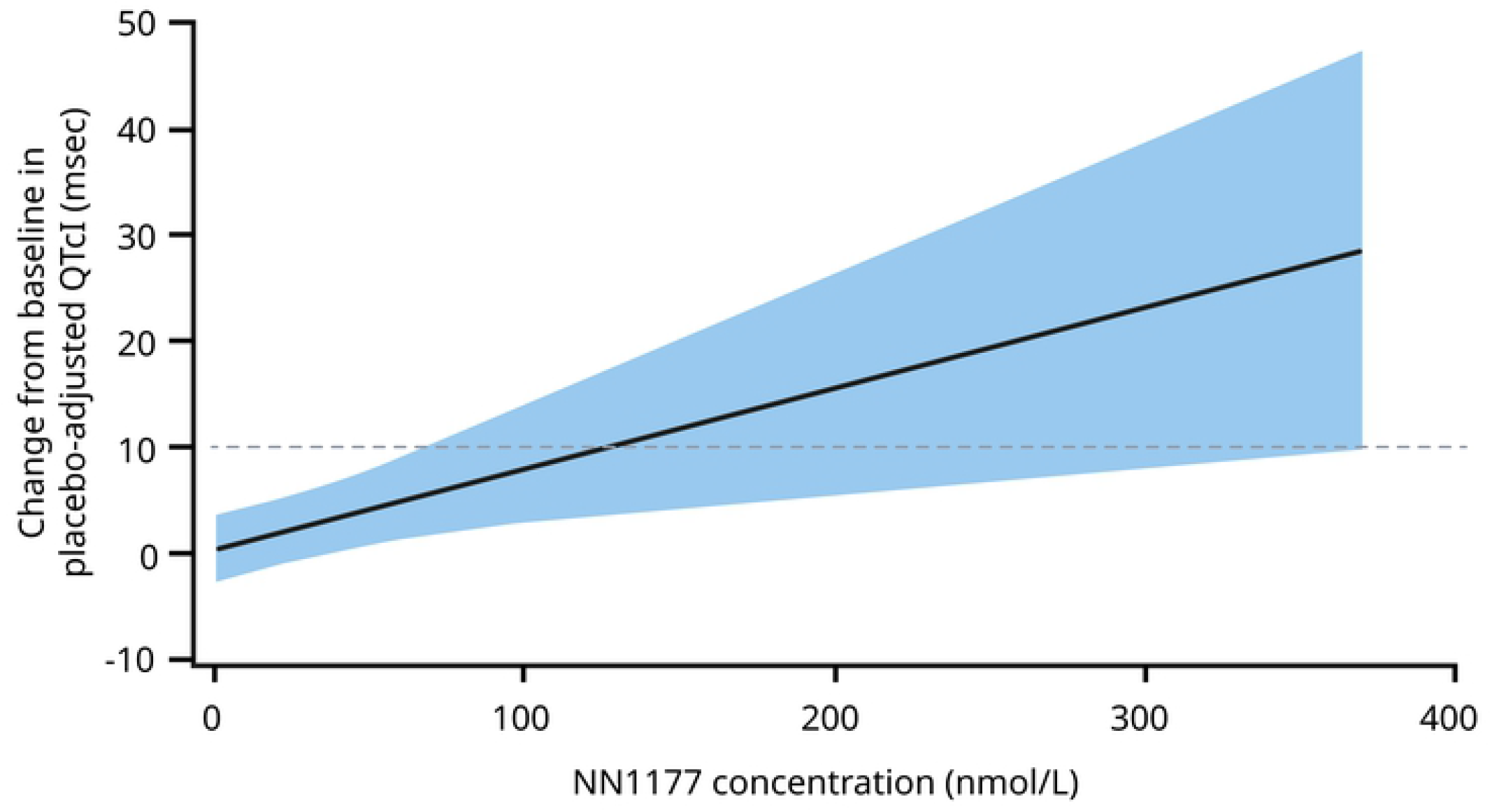
Heart rate in the multiple ascending dose trial (A), mean heart rate by time of day in the drug-drug interaction trial (B), and predicted placebo-adjusted QTcI data change from baseline (C) Data from safety analysis set consisting of all participants who were exposed to at least 1 dose of trial product. A. Vertical reference lines represent first and last dosing of NN1177 B. Vertical reference lines represent first and last dosing of NN1177. Legend shows 24 hours and 6-hour increments. Mean of Holter heart rate during 48 hours for Visit 2 (day-2 to day 1), Visit 2 (day 8 to day 10), Visit 4 (day 22 to day 24), Visit 6 (day 36 to day 38), Visit 8 (day 50 to day 52) and Visit 10 (day 64 to day 66) are plotted on 0, 9, 23, 37, 51 and 65 days, respectively. C. Reference line represents 10 ms prolongation of QTcI. The solid black line with shaded area denotes the model-predicted mean placebo-adjusted delta QTcI with 90% CI. Bpm, beats per minute; CI, confidence interval, DDI, drug-drug interaction; QTcI, QT interval correction using individualised formula.

#### Clinical laboratory analyses

Circulating levels of most amino acids decreased over the course of the DDI trial (Table 4). In the majority of participants, amino acid levels decreased below the lower limit of normal at the last day of dosing (day 78). In the MAD and DDI trials, decreased reticulocyte levels were observed at end of treatment (vs baseline) and, in the MAD trial, the decreases appeared to be dose-dependent in participants treated with NN1177 (≥1,300 μg doses) (Table 5); in both trials, levels had recovered by the end of the follow-up period. There were no clinically relevant changes in other haematological parameters (e.g. erythrocytes, haemoglobin, leucocytes and thrombocytes) in either trial (Table 5 and Supplementary Table S8). In the DDI trial, mean warfarin international normalised ratio (INR) increased following administration of the Cooperstown 5+1 index cocktail; in contrast, no effect of NN1177 was observed on activated partial thromboplastin time (aPTT), prothrombin time INR or albumin levels in the MAD trial (Supplementary Table S8).

**Table 4.**
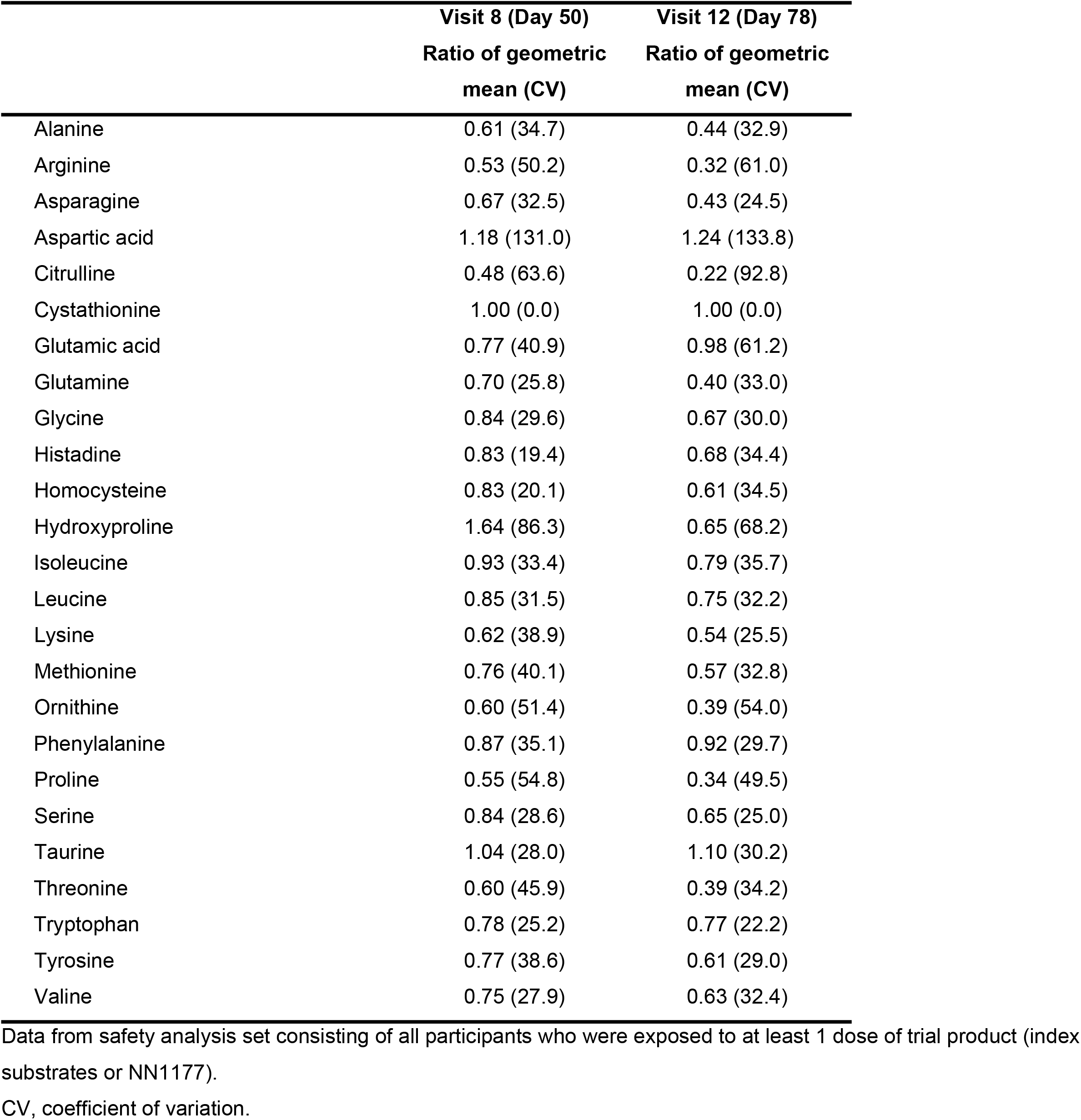
Ratio to baseline for amino acids in the drug–drug interaction trial.

|  | Visit 8 (Day 50) | Visit 12 (Day 78) |
| --- | --- | --- |
|  | Ratio of geometric<br>mean (CV) | Ratio of geometric<br>mean (CV) |
| Alanine | 0.61 (34.7) | 0.44 (32.9) |
| Arginine | 0.53 (50.2) | 0.32 (61.0) |
| Asparagine | 0.67 (32.5) | 0.43 (24.5) |
| Aspartic acid | 1.18 (131.0) | 1.24 (133.8) |
| Citrulline | 0.48 (63.6) | 0.22 (92.8) |
| Cystathionine | 1.00 (0.0) | 1.00 (0.0) |
| Glutamic acid | 0.77 (40.9) | 0.98 (61.2) |
| Glutamine | 0.70 (25.8) | 0.40 (33.0) |
| Glycine | 0.84 (29.6) | 0.67 (30.0) |
| Histadine | 0.83 (19.4) | 0.68 (34.4) |
| Homocysteine | 0.83 (20.1) | 0.61 (34.5) |
| Hydroxyproline | 1.64 (86.3) | 0.65 (68.2) |
| Isoleucine | 0.93 (33.4) | 0.79 (35.7) |
| Leucine | 0.85 (31.5) | 0.75 (32.2) |
| Lysine | 0.62 (38.9) | 0.54 (25.5) |
| Methionine | 0.76 (40.1) | 0.57 (32.8) |
| Ornithine | 0.60 (51.4) | 0.39 (54.0) |
| Phenylalanine | 0.87 (35.1) | 0.92 (29.7) |
| Proline | 0.55 (54.8) | 0.34 (49.5) |
| Serine | 0.84 (28.6) | 0.65 (25.0) |
| Taurine | 1.04 (28.0) | 1.10 (30.2) |
| Threonine | 0.60 (45.9) | 0.39 (34.2) |
| Tryptophan | 0.78 (25.2) | 0.77 (22.2) |
| Tyrosine | 0.77 (38.6) | 0.61 (29.0) |
| Valine | 0.75 (27.9) | 0.63 (32.4) |

Data from safety analysis set consisting of all participants who were exposed to at least 1 dose of trial product (index substrates or NN1177).
CV, coefficient of variation.

**Table 5.** Clinical laboratory parameters in the multiple ascending dose trial.

|  | **NN1177**<br>**200 µg** | **NN1177**<br>**600 µg** | **NN1177**<br>**1,300 µg** | **NN1177**<br>**1,900 µg** | **NN1177**<br>**2,800 µg** | **NN1177**<br>**4,200 µg** | **NN1177**<br>**6,000 µg** | **Placebo** |
| --- | --- | --- | --- | --- | --- | --- | --- | --- |
| **Haematology** |  |  |  |  |  |  |  |  |
| **Haemoglobin (mmol/L)** |  |  |  |  |  |  |  |  |
| Baseline | 8.5 (12.8) | 8.9 (13.6) | 8.3 (12.5) | 8.9 (8.3) | 8.6 (9.0) | 8.2 (10.8) | 8.3 (12.2) | 8.4 (13.1) |
| Ratio to baseline at EOT | 0.95 (3.5) | 0.97 (3.0) | 0.95 (6.5) | 0.94 (8.4) | 0.93 (4.7) | 0.97 (7.7) | 0.93 (2.6) | 0.96 (5.1) |
| Ratio to baseline at follow-up | 1.00 (5.0) | 0.94 (4.6) | 0.94 (2.9) | 0.94 (6.6) | 0.90 (6.0) | 0.94 (6.7) | 0.92 (6.4) | 0.97 (6.1) |
| **Erythrocytes, 10<sup>12</sup>/L** |  |  |  |  |  |  |  |  |
| Baseline | 4.88 (10.6) | 5.08 (13.5) | 4.80 (14.6) | 4.86 (10.3) | 4.62 (9.0) | 4.56 (9.7) | 4.57 (9.5) | 4.77 (11.1) |
| Ratio to baseline at EOT | 0.96 (3.1) | 0.96 (3.4) | 0.96 (7.1) | 0.95 (8.7) | 0.95 (5.5) | 0.98 (8.0) | 0.96 (1.2) | 0.97 (5.2) |
| Ratio to baseline at follow-up | 1.00 (5.4) | 0.94 (5.6) | 0.94 (3.4) | 0.93 (6.0) | 0.91 (5.8) | 0.95 (6.6) | 0.93 (5.3) | 0.97 (6.5) |
| **Reticulocytes, 10<sup>9</sup>/L** |  |  |  |  |  |  |  |  |
| Baseline | 64.35 (32.0) | 67.97 (33.9) | 59.10 (26.2) | 62.78 (43.5) | 69.94 (19.7) | 61.69 (24.4) | 68.96 (19.5) | 63.50 (20.2) |
| Ratio to baseline at EOT | 0.89 (22.7) | 0.95 (24.7) | 0.76 (24.0) | 0.70 (17.5) | 0.63 (27.2) | 0.64 (21.2) | 0.62 (20.3) | 0.90 (18.2) |
| Ratio to baseline at follow-up | 1.07 (14.4) | 1.11 (15.6) | 1.03 (15.5) | 1.07 (24.6) | 0.91 (33.1) | 1.12 (17.8) | 1.05 (38.9) | 1.02 (24.5) |
| **Thrombocytes, 10<sup>9</sup>/L** |  |  |  |  |  |  |  |  |
| Baseline | 268 (17.1) | 243 (14.9) | 252 (23.8) | 262 (24.7) | 248 (9.5) | 251 (21.5) | 248 (23.1) | 264 (28.5) |
| Ratio to baseline at EOT | 1.08 (13.1) | 1.03 (16.6) | 0.96 (14.8) | 0.96 (9.9) | 0.94 (9.2) | 0.94 (18.0) | 1.04 (14.5) | 1.08 (12.6) |
| Ratio to baseline at follow-up | 1.04 (9.8) | 0.97 (15.9) | 1.05 (7.7) | 0.94 (12.5) | 1.02 (9.1) | 1.07 (8.8) | 1.10 (11.6) | 0.99 (13.9) |
| **Leucocytes, 10<sup>9</sup>/L** |  |  |  |  |  |  |  |  |
| Baseline | 6.1 (36.4) | 6.6 (23.9) | 4.9 (22.1) | 5.8 (19.3) | 5.4 (13.2) | 5.8 (33.1) | 6.8 (22.1) | 6.4 (26.6) |
| Ratio to baseline at EOT | 1.04 (19.8) | 1.14 (34.1) | 1.11 (21.9) | 0.97 (16.1) | 0.87 (15.4) | 0.89 (12.9) | 0.85 (17.8) | 1.06 (18.3) |
| Ratio to baseline at follow-up | 1.02 (21.3) | 0.98 (21.2) | 1.15 (17.4) | 1.01 (23.7) | 0.97 (17.8) | 1.03 (10.1) | 0.94 (17.6) | 1.03 (24.2) |
| **Inflammatory biomarkers** |  |  |  |  |  |  |  |  |
| **Fibrinogen, g/L** |  |  |  |  |  |  |  |  |
| Baseline | 2.86 (26.2) | 3.09 (19.7) | 3.37 (15.8) | 2.91 (20.3) | 2.92 (18.5) | 3.20 (15.4) | 3.72 (15.8) | 3.21 (24.9) |
| Ratio to baseline at EOT | 1.01 (16.7) | 1.05 (11.7) | 1.15 (13.9) | 1.14 (0.98) | 1.28 (12.6) | 1.33 (16.0) | 1.36 (12.9) | 0.94 (15.2) |
| Ratio to baseline at follow-up | 0.98 (16.6) | 0.95 (13.6) | 0.89 (10.6) | 0.86 (14.2) | 0.88 (13.0) | 0.98 (24.6) | 0.96 (9.0) | 0.93 (19.1) |
| **hsCRP, mg/L** |  |  |  |  |  |  |  |  |
| Baseline | 1.1 (218.7) | 1.7 (247.7) | 2.9 (109.4) | 1.4 (132.1) | 1.3 (146.1) | 2.0 (146.3) | 2.6 (155.7) | 2.3 (172.0) |
| Ratio to baseline at EOT | 1.22 (93.4) | 0.84 (64.0) | 0.76 (87.4) | 1.11 (82.1) | 1.64 (79.5) | 1.65 (35.6) | 2.14 (99.4) | 1.01 (116.4) |
| Ratio to baseline at follow-up | 1.45 (56.0) | 0.89 (32.4) | 0.90 (38.5) | 0.80 (106.5) | 0.55 (94.8) | 1.18 (106.1) | 1.17 (76.3) | 0.95 (224.2) |
| **Liver parameters** |  |  |  |  |  |  |  |  |
| **Alanine aminotransferase (U/L)** |  |  |  |  |  |  |  |  |
| Baseline | 22 (45.1) | 15 (34.8) | 19 (76.4) | 15 (52.9) | 19 (54.2) | 15 (54.1) | 14 (63.7) | 18 (64.1) |
| Ratio to baseline at EOT | 1.02 (51.8) | 0.97 (11.0) | 1.07 (39.7) | 1.25 (42.3) | 0.93 (65.7) | 1.40 (22.0) | 1.62 (41.2) | 1.03 (39.6) |
| Ratio to baseline at follow-up | 0.82 (55.5) | 1.38 (37.7) | 0.91 (26.0) | 1.19 (46.5) | 1.20 (62.8) | 1.20 (45.1) | 1.56 (56.0) | 0.89 (41.4) |
| **Aspartate aminotransferase (U/L)** |  |  |  |  |  |  |  |  |
| Baseline | 17 (23.7) | 16 (16.7) | 18 (28.6) | 16 (32.4) | 17 (27.6) | 15 (26.8) | 15 (21.3) | 17 (36.7) |
| Ratio to baseline at EOT | 0.94 (28.6) | 1.01 (20.3) | 1.02 (23.3) | 1.02 (29.1) | 0.88 (24.0) | 1.10 (19.0) | 1.25 (19.5) | 1.00 (22.8) |
| Ratio to baseline at follow-up | 0.91 (28.9) | 1.27 (33.2) | 0.88 (20.7) | 1.11 (22.1) | 1.06 (43.5) | 1.05 (27.7) | 1.11 (24.9) | 1.05 (36.0) |

Data from safety analysis set consisting of all participants who were exposed to at least 1 dose of trial product. Baseline information is defined as the measurement at the latest assessment before first dosing. All values are geometric mean (CV).
CV, coefficient of variation; EOT, end of treatment; hsCRP, high-sensitivity C-reactive protein.

In the MAD trial, treatment with NN1177 was associated with increased levels of C-reactive protein (CRP; high-sensitivity assay) and fibrinogen, particularly with the higher dose levels (Table 5). An increase in fibrinogen levels was also seen in the DDI trial. Alanine aminotransferase (ALT) and aspartate aminotransferase (AST) increased from baseline to end of treatment, most apparently in participants receiving the two of the three highest NNC1177 doses (2,800 μg and 4,200 μg; Table 5). However, there were no noteworthy changes in bilirubin or alkaline phosphatase (ALP) levels, or for liver function tests (albumin, aPTT) (Supplementary Table S8). Blood potassium levels decreased in participants treated with NN1177 in both the MAD and DDI trials, and blood sodium levels decreased in the DDI trial (Supplementary Table S8).

### Pharmacodynamics

#### Body weight

In the MAD trial, body weight decreased from baseline through end of treatment (day 85) with NN1177 in a dose-dependent manner (Figure 3).

**Figure 3:**
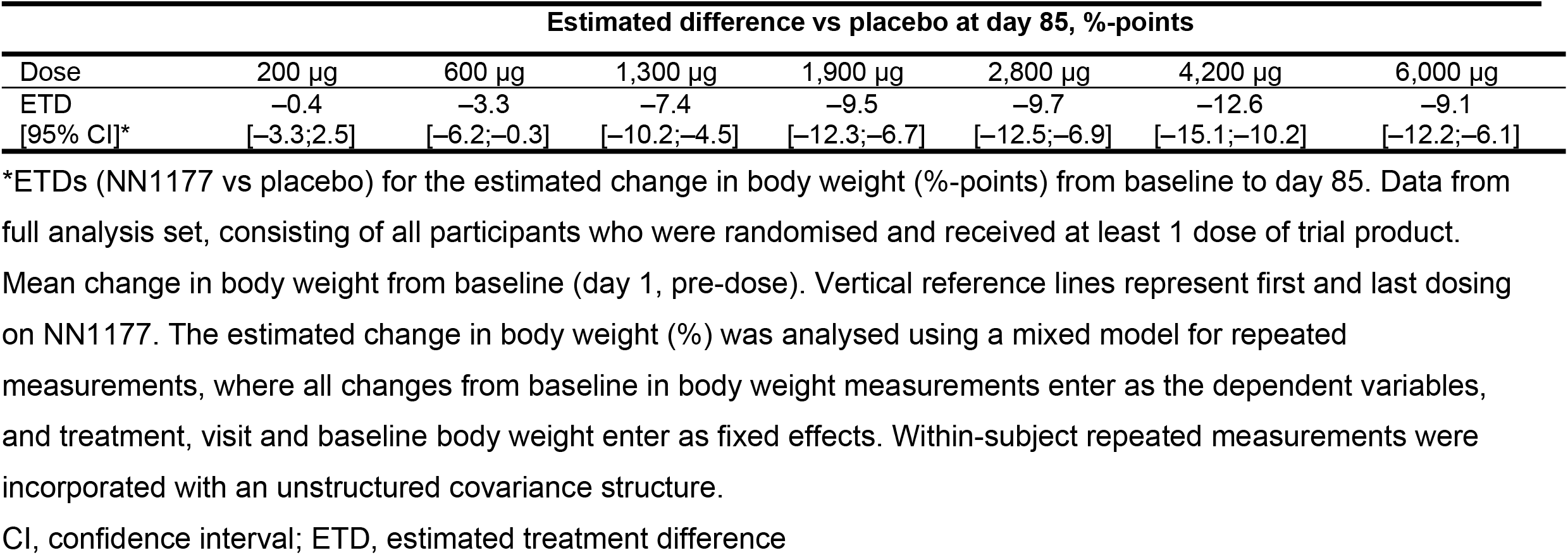
Change in body weight (%) over time in the multiple ascending dose trial for NN1177 and placebo.

The placebo-adjusted estimated body weight changes ranged from 0 kg (200 μg dose) to –12.1 kg (4,200 μg dose) and were statistically significant for the dose levels higher than 200 μg (p<0.05). Following cessation of NN1177 during follow-up, body weight increased in all groups. In the FHD/SAD trial, treatment with NN1177 resulted in weight loss of up to 2.9% (1,100 μg dose; baseline to day 7 post-dosing) and a loss of 10.1% in the DDI trial (baseline to end of treatment), respectively.

#### Leptin

Fasting leptin levels in the MAD trial decreased statistically significantly (vs placebo; p<0.05) with NN1177 at dose levels 1,300 μg and above but with no clear dose dependency; fasting soluble leptin receptors increased statistically significantly (vs placebo; p<0.05) with NN1177 at dose levels 1,900 μg and above in an apparently dose-dependent manner (Supplementary Table S10).

#### Glucose metabolism

In the MAD trial, fluctuations in fasting plasma glucose were observed throughout the treatment period with no clear dose dependency (Figure 4A). Similarly, statistical analysis did not show a clear dose-dependent treatment difference at end-of-treatment. Fluctuations in fasting insulin were observed throughout the treatment period with no clear dose dependency, but a clear increase was observed for the 6,000 ug dose group at end-of-treatment with a statistically significant treatment ratio, compared with placebo, of 2.02 (Figure 4B). During the OGTT, a dose-dependent increase at end-of-treatment was observed for the area under the glucose concentration–time curve from time 0 to 2 hours (AUC_0-2h,glucose_) and in the estimated mean change of 2-hour glucose for the three highest N1177 dose groups, compared with the placebo group (Table 6). Similarly, a dose-dependent increase was observed for the area under the insulin concentration–time curve from time 0 to 2 hours (AUC_0-2h,insulin_). The increase was most apparent for the three highest dose groups (2,800 μg, 4,200 μg and 6,000 μg). There were no clinically relevant changes in HbA_1c_ at end-of-treatment across treatment groups (Figure 4C). In contrast, fasting glucagon decreased from baseline to the follow-up visit for all NN1177 groups (Figure 4D).

**Figure 4.**
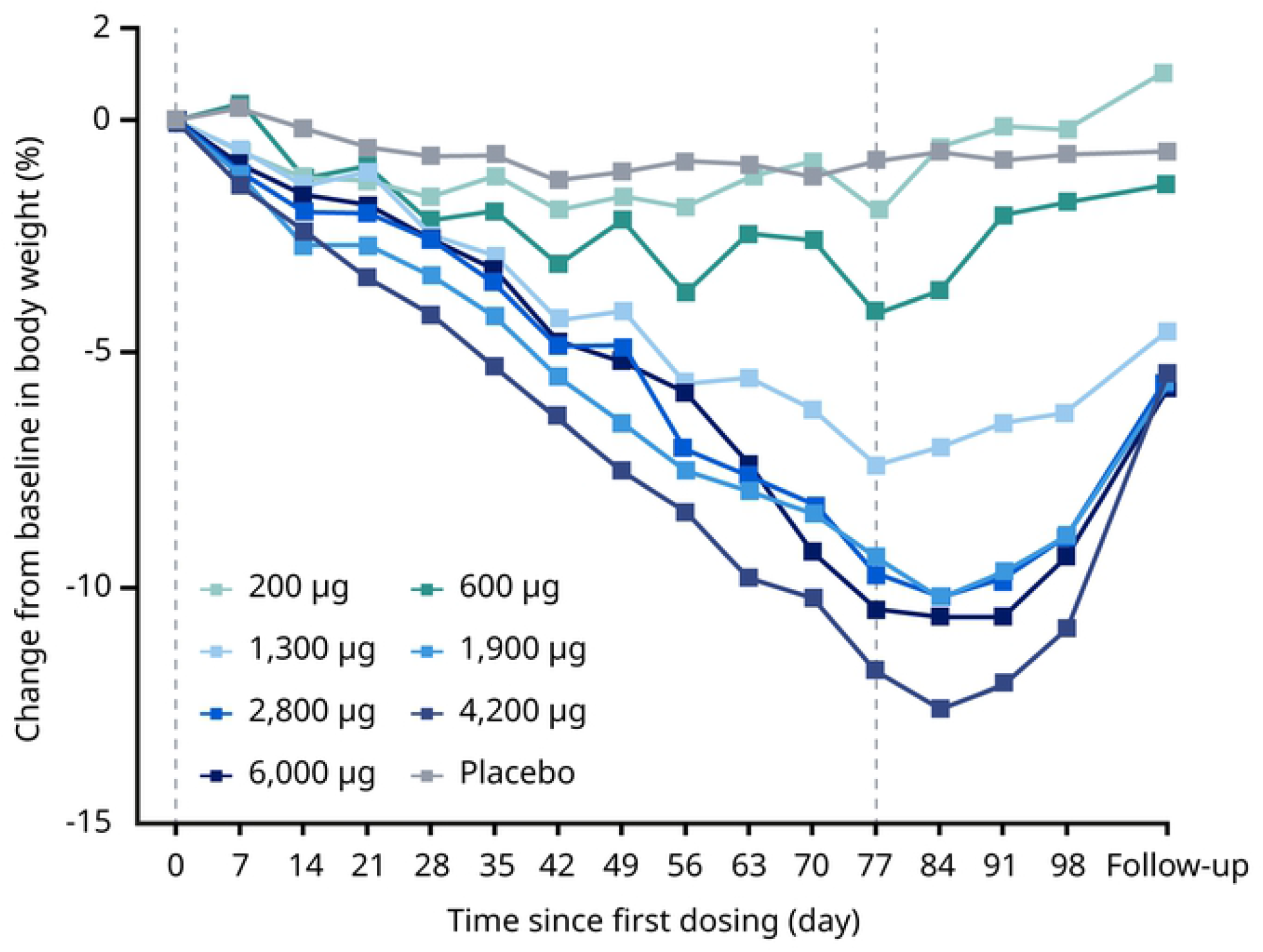
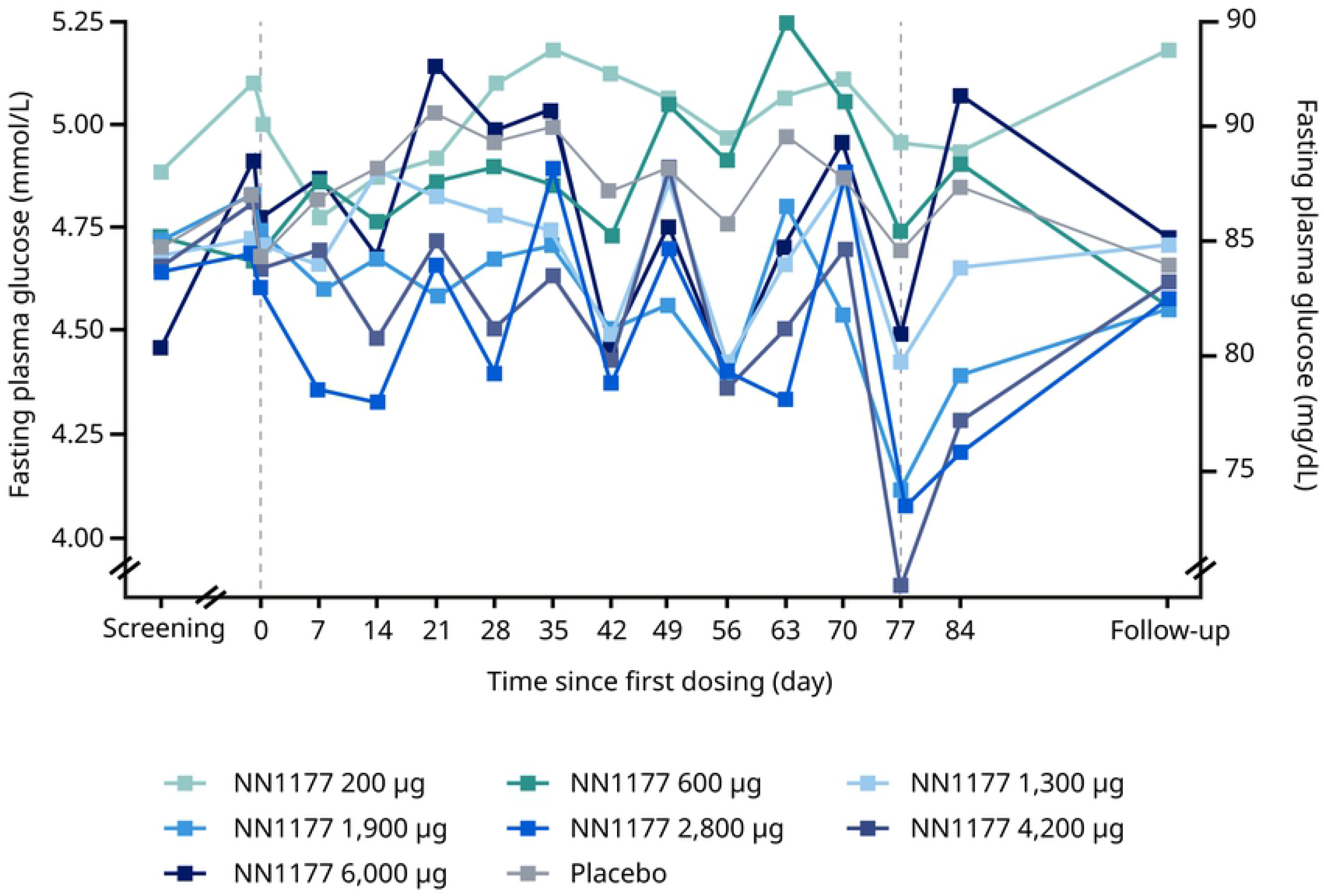
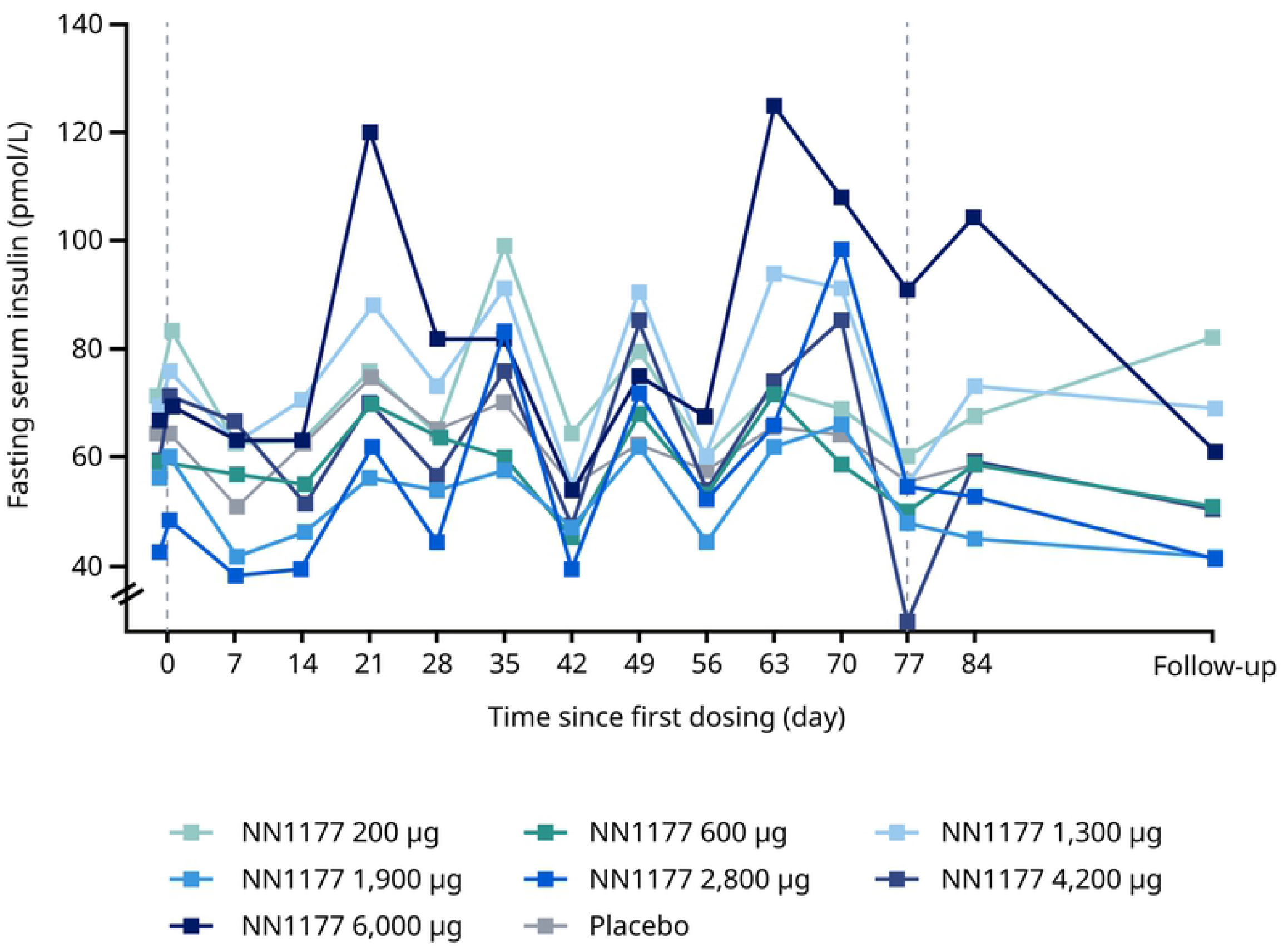
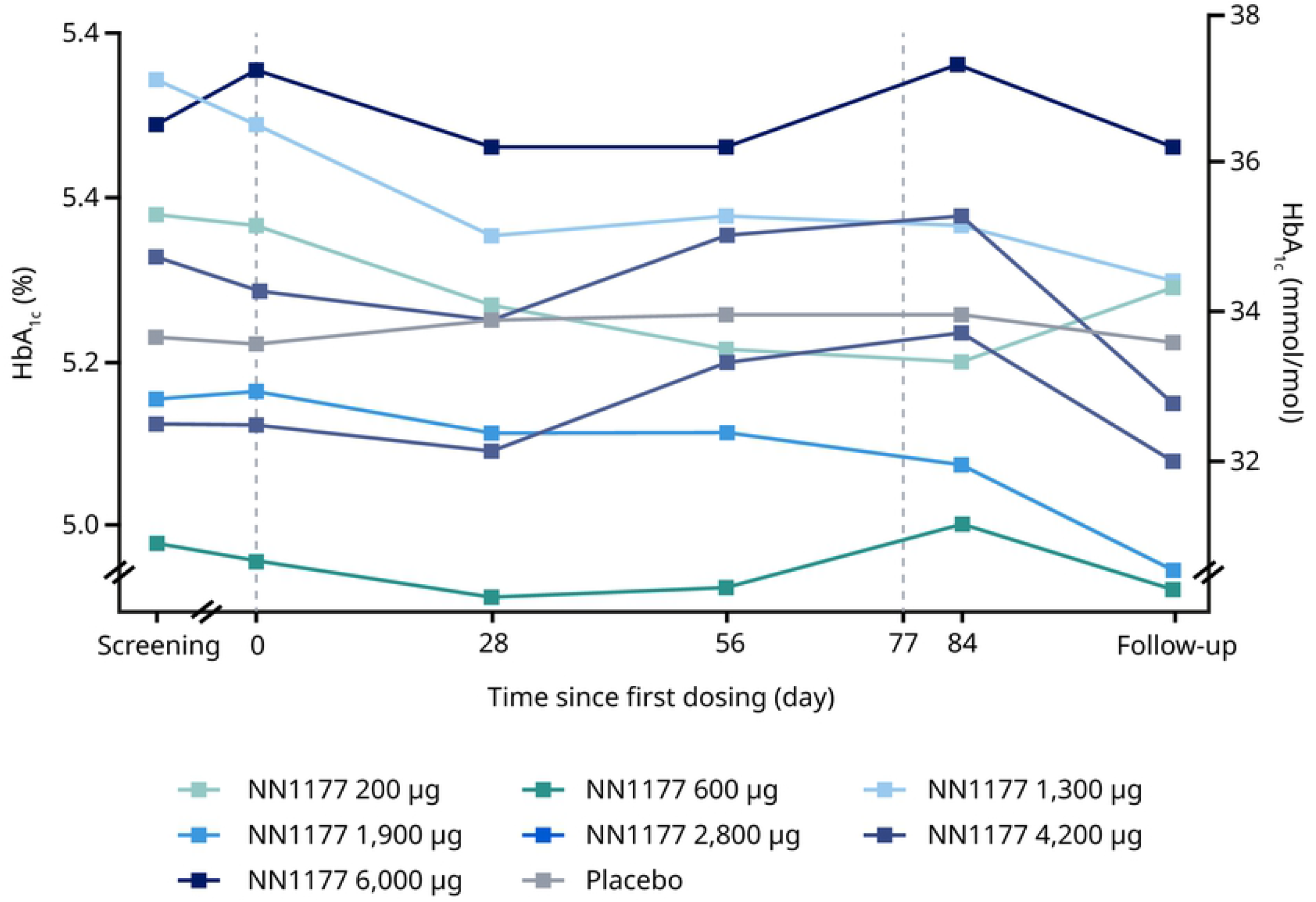
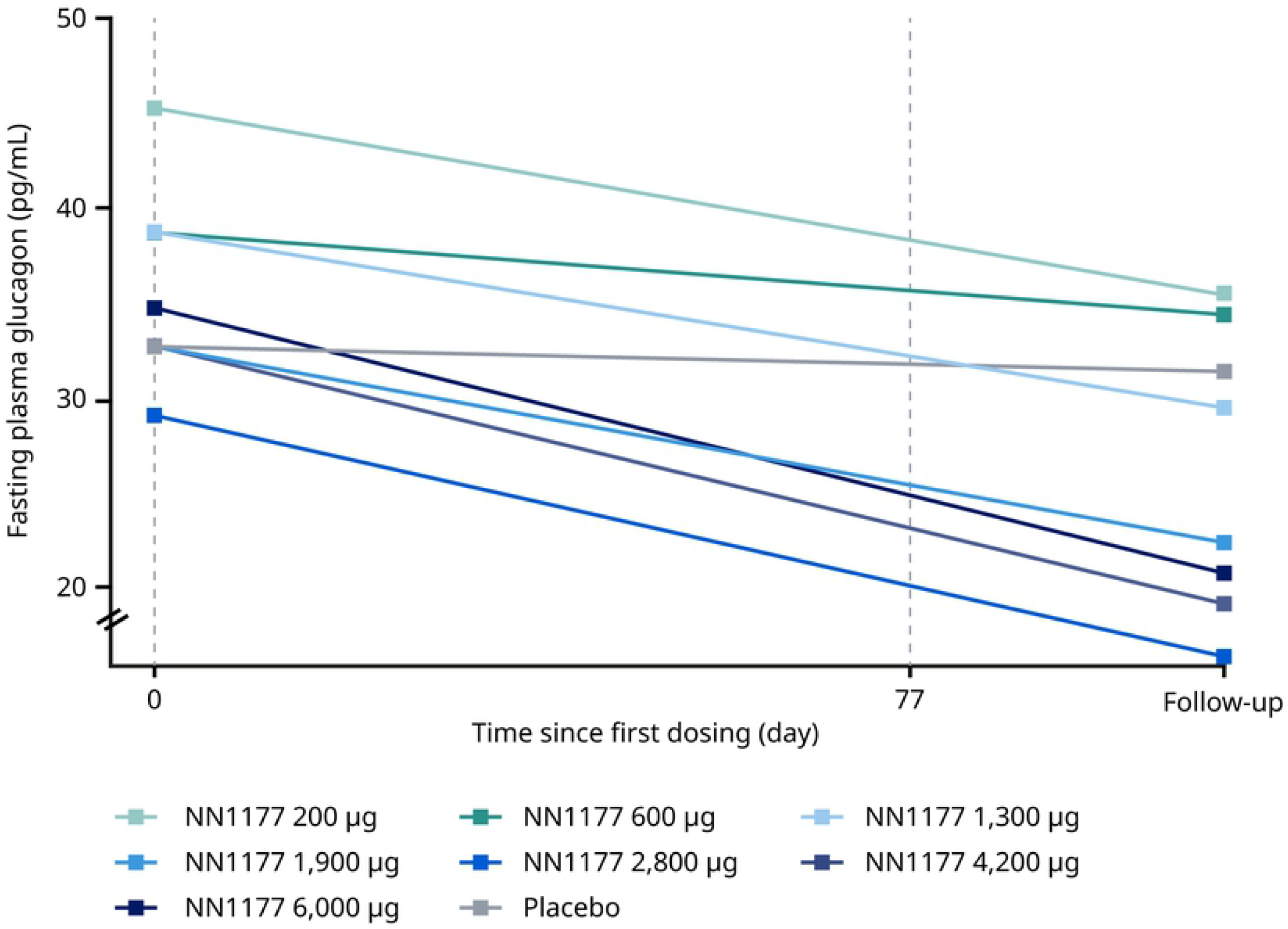
Glucose metabolism endpoints in the multiple ascending dose trial. **A. Fasting glucose** **B. Fasting insulin** **C. Mean HbA**_**1c**_ **D. Fasting glucagon** Data from full analysis set consisting of all participants who were randomised and received at least 1 dose of trial product. Vertical reference lines represent first and last dosing on NN1177 A. Mean fasting glucose from baseline (day –1, pre-dose) to the follow-up visit. B. Geometric mean fasting insulin from baseline (day 1, pre-dose) to the follow-up visit. C. Geometric mean fasting glucagon from baseline, where baseline is defined as the measurement at the latest assessment before dosing. HbA_1c_, haemoglobin A_1c_

**Table 6.**
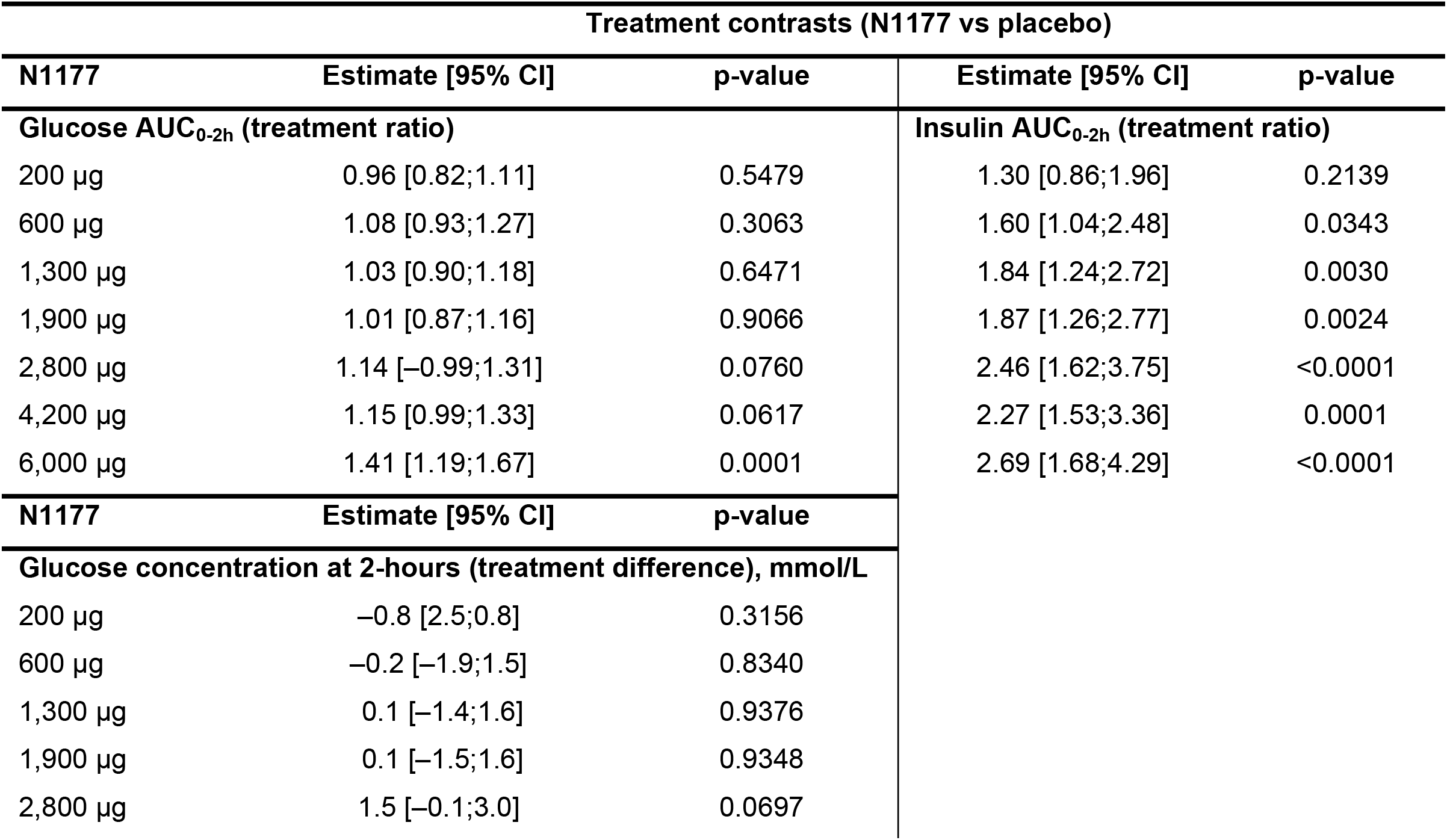
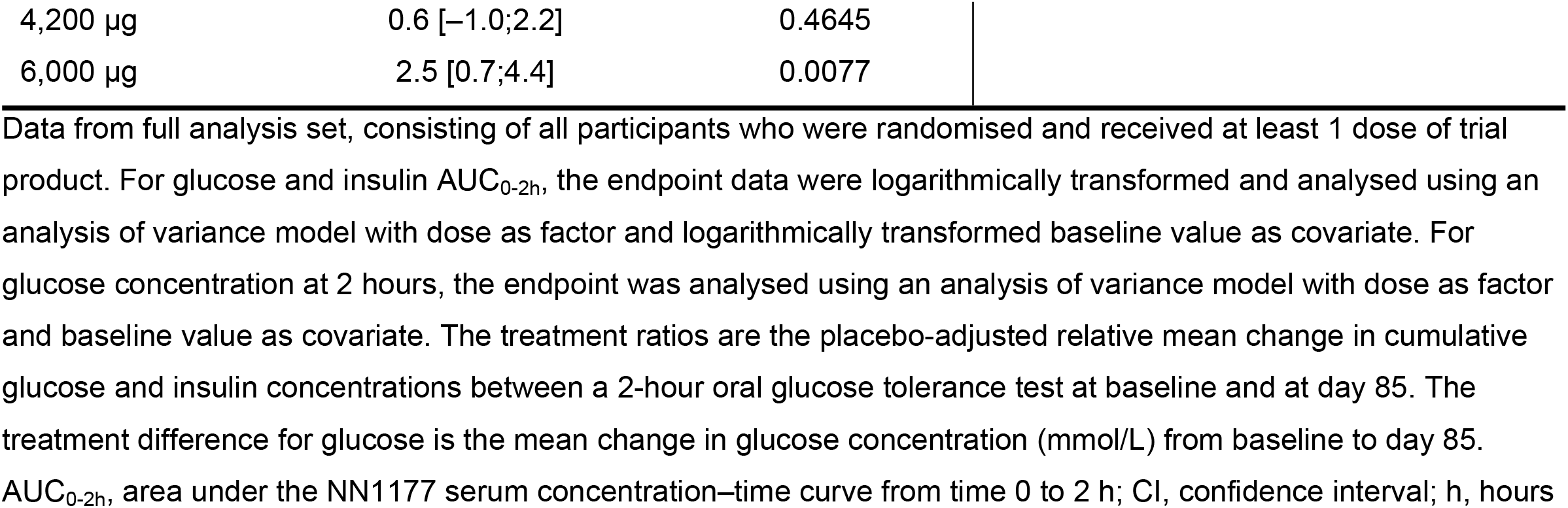
Oral glucose tolerance test in the multiple ascending dose trial.

| Treatment contrasts (N1177 vs placebo) |  |  |  |  |
| --- | --- | --- | --- | --- |
| N1177 | Estimate [95% CI] | p-value | Estimate [95% CI] | p-value |
| **Glucose AUC<sub>0-2h</sub> (treatment ratio)** |  |  | **Insulin AUC<sub>0-2h</sub> (treatment ratio)** |  |
| 200 µg | 0.96 [0.82;1.11] | 0.5479 | 1.30 [0.86;1.96] | 0.2139 |
| 600 µg | 1.08 [0.93;1.27] | 0.3063 | 1.60 [1.04;2.48] | 0.0343 |
| 1,300 µg | 1.03 [0.90;1.18] | 0.6471 | 1.84 [1.24;2.72] | 0.0030 |
| 1,900 µg | 1.01 [0.87;1.16] | 0.9066 | 1.87 [1.26;2.77] | 0.0024 |
| 2,800 µg | 1.14 [–0.99;1.31] | 0.0760 | 2.46 [1.62;3.75] | <0.0001 |
| 4,200 µg | 1.15 [0.99;1.33] | 0.0617 | 2.27 [1.53;3.36] | 0.0001 |
| 6,000 µg | 1.41 [1.19;1.67] | 0.0001 | 2.69 [1.68;4.29] | <0.0001 |
| N1177 | Estimate [95% CI] | p-value |  |  |
| **Glucose concentration at 2-hours (treatment difference), mmol/L** |  |  |  |  |
| 200 µg | –0.8 [2.5;0.8] | 0.3156 |  |  |
| 600 µg | –0.2 [–1.9;1.5] | 0.8340 |  |  |
| 1,300 µg | 0.1 [–1.4;1.6] | 0.9376 |  |  |
| 1,900 µg | 0.1 [–1.5;1.6] | 0.9348 |  |  |
| 2,800 µg | 1.5 [–0.1;3.0] | 0.0697 |  |  |

|  |  |  |
| --- | --- | --- |
| 4,200 µg | 0.6 [-1.0;2.2] | 0.4645 |
| 6,000 µg | 2.5 [0.7;4.4] | 0.0077 |

Data from full analysis set, consisting of all participants who were randomised and received at least 1 dose of trial product. For glucose and insulin AUC<sub>0-2h</sub>, the endpoint data were logarithmically transformed and analysed using an analysis of variance model with dose as factor and logarithmically transformed baseline value as covariate. For glucose concentration at 2 hours, the endpoint was analysed using an analysis of variance model with dose as factor and baseline value as covariate. The treatment ratios are the placebo-adjusted relative mean change in cumulative glucose and insulin concentrations between a 2-hour oral glucose tolerance test at baseline and at day 85. The treatment difference for glucose is the mean change in glucose concentration (mmol/L) from baseline to day 85. AUC<sub>0-2h</sub>, area under the NN1177 serum concentration–time curve from time 0 to 2 h; CI, confidence interval; h, hours

## Discussion

Although treatment with NN1177 in the trials reported herein led to dose-dependent significant weight loss compared with placebo, these were not accompanied by the expected improvements in obesity risk parameters such as glucose metabolism, blood pressure and liver enzymes. Furthermore, several cardiovascular and general safety concerns were raised for the higher NN1177 doses in the MAD trial, which prompted a decision to discontinue clinical development of the glucagon/GLP-1 receptor co-agonist. Indeed, preclinical evaluation of NN1177 revealed a complex picture of variability of compound exposure and study length when determining the optimal receptor balance [27]. This may partly explain the impeded clinical success. However, our results for NN1177 are consistent with other studies demonstrating weight loss and related beneficial outcomes for glucagon and GLP-1 RAs [2, 9, 16, 17].

A lower than expected estimated placebo-adjusted weight loss was observed with NN1177 at the maximum 6,000 μg dose (9.1%) compared with the 4,200 μg dose (12.6%); this observation may have partly been due to the modified dose escalation. However, more importantly, weight loss with NN1177 was also accompanied by adverse tolerability and safety findings, including an increased prevalence of GI AEs, which appeared to be dose-related from 700 μg.

Additionally, increased heart rate (including blunted night-time dip) was observed with NN1177. Indeed, GLP-1 RAs are known to have chronotropic effects; for example, increases in pulse rate have been reported in participants with type 2 diabetes in a MAD trial with semaglutide [31, 32] and in trials with dulaglutide and a dual glucose-dependent insulinotropic polypeptide (GIP) and GLP-1 RA [33]. However, the increase in heart rate for NN1177 far exceeds that observed for semaglutide; for example in the SUSTAIN 6 cardiovascular outcomes trial for semaglutide in type 2 diabetes, a placebo-corrected heart rate increase of 3.2 bpm was reported for the 1.0 mg dose level [34] and in STEP 1 for patients with overweight or obesity, mean increases in resting heart rate of 1 to 4 bpm were observed for semaglutide compared to placebo [35]. Furthermore, the increase in heart rate in the MAD and DDI trials, together with the blunting of the nocturnal dip in heart rate, may be detrimental, because a study has shown that blunted nocturnal heart rate dipping is associated with preclinical cardiac damage (left atrial enlargement) and is predictive of cardiovascular morbidity and mortality in the general population [36]. It is unknown if the heart rate increase would be sustained for longer periods than tested in the trials, or if NN1177 tachyphylaxis would lead to heart rate normalisation over time. However, it was concluded that the magnitude of the heart rate increase observed with NN1177 at clinically relevant doses was not acceptable.

Analysis of electrocardiograms in the MAD trial showed a statistically significant increase in QTcI with clinically relevant doses of NN1177. QTcI prolongation may be a glucagon effect, since QT prolongation has not been observed for receptor agonists of GLP-1 alone [37, 38]. However, the concentration–QTc slope estimate for NN1177 in the MAD trial should be interpreted with caution because the hysteresis plot in Supplementary Figure S3B indicates a potential time delay between concentration and QT.

The clinical laboratory analyses also revealed some potentially relevant adverse effects of NN1177 treatment. There were marked reductions in serum levels of most amino acids in the DDI trial. Reductions in plasma amino acid levels have previously been observed with glucagon treatment [39] and may indicate that amino acid homeostasis is disturbed, potentially reflecting increased hepatic gluconeogenesis (through glucagon stimulation) with amino acids as substrate. Indeed, such amino acid reductions are not seen at similar magnitude following bariatric surgery [40]. Substantial decreases in circulating amino acids have been observed in patients with glucagonoma, where there is excessive production of glucagon [12, 13]. In patients with critical illness, elevated plasma glucagon promotes gluconeogenesis, which requires skeletal muscle amino acid utilisation, ultimately leading to muscle wasting and depressed blood amino acid levels [41]. Studies have shown that people with elevated circulating concentrations of glucagon over 7-fold higher than normal are at risk of protein wasting, hypoaminoacidaemia and weight loss [42]. Similarly, necrolytic migratory erythema, which is also seen in glucagonoma [43] and with prolonged treatment for congenital hyperinsulinaemia [44], may be linked with a decrease in circulating amino acids [45]. Therefore, because it is unknown whether a decrease in circulating amino acids will increase protein breakdown in lean tissue, the clinical consequences that could arise from the reduction of amino acids seen with NN1177 are currently unclear.

In the DDI trial, decreases in reticulocyte levels were observed and may indicate bone marrow suppression [46]; however, although a direct inhibitory effect of glucagon on erythropoiesis has been observed [47], evidence is sparse. Reduced food and micronutrient intake and catabolic state may also contribute to the reduction in reticulocytes observed in our trials. Increases in INR were seen when NN1177 and warfarin were co-administered, despite administration of vitamin K. As noted in the full report of the DDI trial [28], the increase in mean INR is not explained by warfarin PK. Although the interaction between glucagon and warfarin has been described previously, it is not fully understood [48].

Fibrinogen and CRP levels also increased in the trials, pointing to a possible proinflammatory effect of NN1177. With respect to fibrinogen, the clinical significance of the observed increases is unknown, because there is conflicting evidence for causality between hyperfibrinogenaemia and, for example, cardiovascular disease [49]. Because there is limited evidence of a direct stimulatory effect of glucagon on fibrinogen synthesis [50], our findings could be the result of a potential acute phase response caused by metabolic stress or hepatic stimulation. In addition, increased levels of liver parameters (AST and ALT) were noted in our trials; however, there were no concerning cases in participants in the trials, and no clear evidence of hepatocellular injury. Other studies suggest that some liver enzymes may transiently increase during a dietary-induced weight loss [51, 52]. In one study, this effect varied according to sex [22]; however, overall, the effect is considered to be due to a transient deterioration in hepatic steatosis prior to improvement and, therefore, a benign effect [51, 52]. In contrast, a previous phase 2 trial with semaglutide in participants with obesity showed decreases in liver enzymes (ALT) and CRP [53]. Indeed, GLP-1 RAs like liraglutide and semaglutide are known to reduce inflammation as measured by CRP [54].

Although the trial population did not have type 2 diabetes, the effects of NN1177 on body weight in the MAD and DDI studies had no beneficial effect on glycaemic parameters. Of concern is the impaired glucose tolerance observed during the OGTT despite clinically significant weight loss. This result is in contrast to observations with GLP-1 RAs, for which increased glucose tolerance and weight loss is observed in adults with obesity following treatment, for example the STEP 1 trial where placebo-adjusted weight loss was 14.4% at week 68 [55]. Lack of benefit or even detrimental effects on glycaemic parameters would not be well received for weight-loss compounds that are intended to be used in a population where the prevalence of pre-diabetes and type 2 diabetes is high.

## Conclusions

Whilst dose-dependent weight loss was shown with NN1177, the dose-dependent safety concerns observed with up to 12 weeks treatment were considered too pronounced to support the continued clinical development of NN1177, a glucagon/GLP-1 receptor co-agonist for weight management.

## Data Availability

All relevant data are within the manuscript and its Supporting Information files.

## Acknowledgements

We thank all the participants, investigators and trial-site staff who were involved in the conduct of the trials.

We also thank Christoffer Tornøe, Mette Holt, Kirsten Raun and Kevin Tan for their review and input to the manuscript, and Tracey Jones-Hughes (from AXON Communications), for medical writing and editorial assistance (funded by Novo Nordisk).

## Disclosures

M. F., L.E., F.F.K, S.T. and S.B.N. are employees of Novo Nordisk A/S. M.F. and L.E., F.F.K. are shareholders in Novo Nordisk A/S. M.K. has received funding from Diffusion Pharmaceutical Inc., Grifols, Urovant Sciences, ViroDefense, Merck, PhaseBio Pharmaceuticals, Inc., Idorsia Pharmaceuticals Ltd, DynPort Vaccine company/FDA/NIH, and Aerovate Therapeutics. R.G. is an employee of Parexel International.

## Author contributions

All authors contributed to data interpretation and writing the manuscript (reviewing and editing) and approved the final version for publication. The authors had full access to all the data in the studies and were responsible for the decision to submit for publication.

## Funding

Novo Nordisk A./S.

## Supporting information

**S1 Appendix**

**S2 Appendix**

Table S1. Dose escalation regimens and time on target dose for (A) multiple ascending dose and (B) first human dose/single ascending dose trials

A. Based on experience from the first 6 cohorts, dose escalation regimen for cohort 7 was modified. B. Due to gastrointestinal tolerability signals, the planned dose levels of 1,600 and 2,000 μg were not initiated.

Table S2. Baseline characteristics for drug-drug interaction trial

Data from full analysis set consisting of all participants who were randomised and received at least 1 dose of trial product (index substrates or NN1177). Data are means (SD, or range for age) unless otherwise stated)

BMI, body mass index; HbA_1c_, glycated haemoglobin; N, number of participants with available data; NA, not available; pressure; SD, standard deviation

Table S3. Baseline characteristics by dose level in the multiple ascending dose and first human dose/single ascending dose trials

Data were from the safety analysis sets, consisting of all participants who were randomised and received at least 1 dose of trial product. Data are mean unless otherwise specified. Baseline information is defined as the measurement at the latest assessment before dosing. BMI is calculated based on baseline measurements of body weight and height. %, percentage of subjects with characteristic; BMI, body mass index; HbA_1c_, glycated haemoglobin; LDL, low-density lipoprotein; MAD, multiple ascending dose; Max., maximum; Min., minimum; N, number of participants with available data.

Table S4. Pharmacokinetic endpoints in the first human dose/single ascending dose trial

Data from the full analysis sets consisting of participants who were randomised and received at least 1 dose of trial product. Pharmacokinetic endpoints of NN1177 after a single dose. Increases in AUC_0-∞_ and C_max_ with increasing dose were consistent with dose proportionality. The total apparent clearance after a single dose of NN1177 was calculated as CL/F=dose/AUC_0-∞._ The apparent volume of distribution of NN1177 during elimination after a single dose was estimated as V_z_/F=(CL/F)/λ_z_. The terminal rate constant, λ_z_, was estimated by log-linear regression on the terminal part of the concentration–time curve of NN1177 after a single dose using ≥3 observations above lower limit of quantification. The terminal half-life of NN1177 after a single dose was determined as t_½_=log(2)/λ_z_. AUC, area under the curve; C_max_, maximum serum concentration; CL/F, the apparent total serum clearance of NN1177; CV, coefficient of variation; Max., maximum; Min., minimum; MRT, the mean residence time; t_1/2_, time to terminal half-life; t_max_, time to maximum serum concentration; V_z_/F, apparent volume of distribution during elimination

Table S5. Reported treatment-emergent adverse events by preferred term in the multiple ascending dose and first human dose/single ascending dose trials where total events ≥10% Data from safety analysis set, consisting of all participants who were exposed to at least 1 dose of trial product. %, percentage of participants with event(s); E, number of events; N, number of participants with event

Table S6. Reported treatment emergent adverse events in the first human dose/single ascending dose trial

Data from safety analysis set, consisting of all participants who were exposed to at least 1 dose of trial product.

%, percentage of participants with event(s); E, events; n, number of participants with event

Table S7. Reported treatment emergent adverse events in the drug-drug interaction trial

Data from safety analysis set, consisting of all participants who were exposed to at least 1 dose of trial product (index substrates or NN1177). %, percentage of participants with event(s); DDI, drug–drug interaction; E, number of events; N, number of participants with event(s).

Table S8. Clinical laboratory parameters for multiple ascending dose trial

Data from safety analysis set, consisting of all participants who were exposed to at least 1 dose of trial product. Baseline information is defined as the measurement at the latest assessment before dosing. All values are geometric mean (CV).

CV, coefficient of variation in percentage; EOT, end of treatment; hsCRP, high-sensitivity C-reactive protein

Table S9. Clinical laboratory parameters for first human dose/single ascending dose trial

Data from safety analysis set, consisting of all participants who were exposed to at least 1 dose of trial product. Baseline information is defined as the measurement at the latest assessment before dosing. All values are geometric mean (CV).

CV, coefficient of variation in percentage; EOT, end of treatment; hsCRP, high-sensitivity C-reactive protein

Table S10. Fasting leptin for multiple ascending dose trial

Data from full analysis set. Change in fasting leptin was analysed using a mixed model for repeated measurements, where all logarithmically transformed relative changes from baseline in fasting leptin measurements enter as the dependent variables. Treatment, visit and logarithmically transformed baseline fasting leptin enter as fixed effects. Treatment and baseline effect are nested within visit. Within-subject repeated measurements are incorporated with an unstructured covariance structure.

CI, confidence interval

Figure S1. Pharmacokinetic profiles for NN1177 in the multiple ascending dose (A) and first human dose/single ascending dose (B) trials

Data from full analysis sets consisting of participants who were randomised and received at least 1 dose of trial product. Geometric mean plot of NN1177 profiles on logarithmic scale. Values below lower limit of quantification were imputed

Figure S2. Mean QTcI as a function of time for the multiple ascending dose trial

Data from safety analysis set consisting of all participants who were exposed to at least 1 dose of trial product. Vertical reference lines represent first and last dosing of NN1177.

QTcI, QT interval correction using individualised formula.

Figure S3. Model diagnostics plots for QTcI for the multiple ascending dose trial

Data from safety analysis sets consisting of all participants who were exposed to at least 1 dose of trial product. A. QTcI vs RR with model predicted mean (95% CI). B. Plot of mean placebo-adjusted change in QTcI change vs concentration by active treatment. Marker labels display nominal time points of assessments.

CI, confidence interval; QTcI, QT interval correction using individualised formula; RR, time between two successive ventricular depolarisations.

## Notes

### Competing Interest Statement

M. F., L.E., F.F.K, S.T., S.B.N. are employees of Novo Nordisk A/S. M.F., L.E., F.F.K. are shareholders in Novo Nordisk A/S. R.G. is an employee of Parexel International M.K. has received funding from Diffusion Pharmaceutical Inc., Grifols, Urovant Sciences, ViroDefense, Merck, PhaseBio Pharmaceuticals, Inc., Idorsia Pharmaceuticals Ltd, DynPort Vaccine company/FDA/NIH, and Aerovate Therapeutics

### Clinical Trial

ClinicalTrials.gov ID NCT03308721 ClinicalTrials.gov NCT04059367 ClinicalTrials.gov ID NCT02941042

### Clinical Protocols

https://clinicaltrials.gov/ct2/show/NCT02941042

https://clinicaltrials.gov/ct2/show/NCT04059367

https://clinicaltrials.gov/ct2/show/NCT03308721

### Funding Statement

This study was funded by Novo Nordisk A/S.

### Author Declarations

The studies obtained approval from local institutional review boards (Aspire IRB 11491 Woodside Ave Santee, CA 92071 and Midlands Independant Review Board 8417 Santa Fe Drive/Suite 100 Overland Park, KS 66212). Written consent was obtained from participants in all three trials.

